# Modelling the epidemiological implications for SARS-CoV-2 of Christmas household bubbles in England

**DOI:** 10.1101/2022.07.04.22277231

**Authors:** Edward M. Hill

## Abstract

The emergence of SARS-CoV-2 saw severe detriments to public health being inflicted by COVID-19 disease throughout 2020. In the lead up to Christmas 2020, the UK Government sought an easement of social restrictions that would permit spending time with others over the Christmas period, whilst limiting the risk of spreading SARS-CoV-2. In November 2020, plans were published to allow individuals to socialise within ‘Christmas bubbles’ with friends and family. This policy involved a planned easing of restrictions in England between 23-27 December 2020, with Christmas bubbles allowing people from up to three households to meet throughout the holiday period. We estimated the epidemiological impact of both this and alternative bubble strategies that allowed extending contacts beyond the immediate household. We used a stochastic individual-based model for a synthetic population of 100,000 households, with demographic and SARS-CoV-2 epidemiological characteristics comparable to England as of November 2020. We evaluated five Christmas bubble scenarios for the period 23-27 December 2020, assuming our populations of households did not have symptomatic infection present and were not in isolation as the eased social restrictions began. Assessment comprised incidence and cumulative infection metrics. We tested the sensitivity of the results to a situation where it was possible for households to be in isolation at the beginning of the Christmas bubble period and also when there was lower adherence to testing, contact tracing and isolation interventions. We found that visiting family and friends over the holiday period for a shorter duration and in smaller groups was less risky than spending the entire five days together. The increases in infection from greater amounts of social mixing disproportionately impacted the eldest. We provide this account as an illustration of a real-time contribution of modelling insights to a scientific advisory group, the Scientific Pandemic Influenza Group on Modelling, Operational sub-group (SPI-M-O) for the Scientific Advisory Group for Emergencies (SAGE) in the UK, during the COVID-19 pandemic. This manuscript was submitted as part of a theme issue on “Modelling COVID-19 and Preparedness for Future Pandemics”.

## 1 Introduction

The COVID-19 pandemic has had grave consequences for global public health. In the initial absence of known effective therapeutics and vaccinations against SARS-CoV-2, the strategies implemented by many nations during 2020 to attempt to curb SARS-CoV-2 outbreaks revolved around nonpharmaceutical interventions (NPIs), including stringent lockdown rules and social distancing measures [1]. Despite the use of such measures in England, by 30 November 2020 there had been 52,881 people who died within 28 days of being identified as a COVID-19 case by a positive test [2].

The vacation period around Christmas is an important time for many people of all faiths and none who come together over the holidays. In light of the hardships suffered during 2020, the UK government was considering actions to balance the desire to spend time with others over the Christmas period, whilst limiting the risk of spreading SARS-CoV-2. On that basis, on 24 November 2020 plans were published to allow individuals to socialise within ‘Christmas bubbles’ with friends and family [3]. This policy involved a planned easing of restrictions in England during 23–27 December 2020. The intent was that anyone who was not self-isolating could form a Christmas bubble according to three main rules: (i) only being in one Christmas bubble; (ii) not changing their Christmas bubble; and (iii) the Christmas bubble could not include people from more than three households. Christmas bubbles could meet, without socially distancing, in private homes, places of worship or public outdoor spaces.

Ultimately, these original plans for Christmas bubbles never came to fruition, being heavily revised following the emergence of the (later named) Alpha variant in Kent, England [4]. On 19 December 2020 [5], it was announced that Christmas bubbles in England would be permitted only on Christmas Day (25 December 2020) and only in locations not in the highest level of restrictions (Tier 4) [6].

Nonetheless, under the proviso of the original plan for Christmas bubbles in England throughout 23-27 December 2020, the expected increase in social interactions during that time meant there was interest in ascertaining the epidemiological implications of household bubbles. Contact clustering in social bubbles beyond the immediate household had already undergone consideration during the COVID-19 pandemic, as part of both modified lockdown policies and gradual lockdown exit strategies granting greater social interaction [7, 8]. It had been demonstrated that social network-based reduction of contact strongly enhanced the effectiveness of social distancing measures while keeping risks lower [7]. Prior work using an individual based model had estimated the impact of such bubble strategies on epidemic and mortality risk using the UK as a case study, finding that social bubbles could be an effective way of extending contacts beyond the household while limiting the increase in epidemic risk, if managed appropriately [8].

The data and science surrounding the SARS-Cov-2 infection is fast moving. In the context of Christmas bubbles in England during 2020, contributory tools to evaluate epidemiological questions on household bubbles and their ramifications included scenario assessments using mathematical models. We present here our associated analyses conducted in late November and early December 2020, demonstrating the potential uses of a stochastic individual-based model representing a synthetic population of households, which could be developed swiftly and deployed in situations necessitating assessment of the epidemiological impact of extending contacts beyond the immediate household. We treat our manuscript as a record of the state of our modelling and the epidemiological context at that time. There have been subsequent additions to the literature relevant to this topic, which are remarked upon in the Discussion.

In this study, we describe a stochastic individual-based model that was used to compare a set of household bubble scenarios during 23-27 December 2020 in England. To enable its prompt investigation the model was bespoke and parsimonious, comprised of a synthetic population of households initialised to reflect the demography and epidemiological context of England, with no symptomatic infection present and with between-household mixing occurring through implementation of a household visit schedule. Purely focused on epidemiological outcomes, we found conditional on there being any easing of rules on household gatherings then shorter and smaller gatherings were preferred. Furthermore, the model quantified the increase in incidence in the over 65s in circumstances where additional mixing occurred over more days.

We present the manuscript as an example of the real-time contribution mathematical epidemiological and modelling of infectious disease dynamics can make to the knowledge base informing policy in response to a public health emergency. An earlier version of this work was presented to the Scientific Pandemic Influenza Group on Modelling, Operational sub-group (SPI-M-O) for the Scientific Advisory Group for Emergencies (SAGE) in the UK on 02 December 2020, contributing to paragraphs in the consensus statement on the impact of a festive break in relaxations [9].

## 2 Methods

We simulated an epidemic process amongst a population of households and assessed the epidemiological impact of temporarily expanding the permitted size of household bubbles during Christmas 2020.

In the Methods we detail: (i) the parameterisation of the households (Section 2.1), (ii) the epidemiological model for SARS-CoV-2 transmission (Section 2.2), (iii) our implementation of test, trace and isolate guidance (Section 2.3), (iv) the Christmas bubble scenarios evaluated (Section 2.4), and (v) the simulation protocol used to assess the Christmas bubble scenarios (Section 2.5).

### 2.1 Household demographics

In our model, each individual was assigned to a household and classified into one of three age groups: 0-19yrs, 20-64yrs, 65+yrs. The three-class age structure allowed for age-based heterogeneity in probabilities of asymptomatic infection and susceptibility to infection. We considered a population of 100,000 households, sampling household sizes and the proportion of households satisfying a given age composition (e.g. one 0-19yrs individual and two 65+ year olds) from 2011 census data for England and Wales [10].

Single-occupancy households and single parent households could form support bubbles with another household [11], which we refer to as an extended household. We modelled extended households as, in essence, having the same transmission rates as if they shared the same physical household over the simulated period.

In the absence of data on the proportion of single person households/single-parent households in a support bubble, for each simulation we sampled the propensity to form a support bubble from a Uniform(0.5,0.75) distribution (*i*.*e*. sampling on the basis of the majority of those households joining up with another).

### 2.2 SARS-CoV-2 epidemiological model

In this section we outline our assumptions regarding the natural course of infection, asymptomatic infection and transmission, susceptibility and the relationship between transmission risk and household size. For a listing of associated parameters, see Table 1.

**Table 1:** Description of epidemiological parameters. The stated distributions are as reported in the cited sources, with additional context provided in the associated subsections of the main text.

| Description | Distribution | Source |
| --- | --- | --- |
| Incubation period | Erlang(6, 0.88) | [12] |
| Infectiousness profile | Infectivity profile over 14 days:<br>[0.0369, 0.0491, 0.0835, 0.1190,<br>0.1439, 0.1497, 0.1354, 0.1076,<br>0.0757, 0.0476, 0.0269, 0.0138,<br>0.0064, 0.0044] | [13, 14] |
| Proportion of cases asymptomatic (0-19yrs) | Uniform(0.20, 0.35) | [15] |
| Proportion of cases asymptomatic (20+yrs) | Uniform(0.05, 0.20) | [15] |
| Relative infectiousness of an asymptomatic | Uniform(0.30, 0.70) | [15, 16] |
| Relative susceptibility of 0-19yrs age class | Uniform(0.40, 0.60) | [17] |
| Household attack rates | Size 2: Normal(0.48,0.06)<br>Size 3: Normal(0.40,0.06)<br>Size 4: Normal(0.33,0.05)<br>Size $\geq 5$: Normal(0.22,0.05) | [18] |

#### 2.2.1 Disease state structure

We ran a disease process on the households, highlighting that we only considered infection resulting from person-to-person interactions due to household mixing. We did not consider transmission arising from other settings based on a wish to evaluate the additional infections that would arise from increased between-household mixing. The implications of this assumption is expanded upon in the Discussion.

We modelled the natural infection history using a Susceptible-Exposed-Infected-Recovered (SEIR) type structure. Namely, each individual was either in a susceptible, exposed/latent (infected but not infectious), infectious (asymptomatic, pre-symptomatic or symptomatic) or recovered state. Once infected, we assumed infectiousness could start from the following day. We assumed an Erlang-distributed incubation period to symptom onset, parameterised using the central parameter estimates for an Erlang-distributed incubation period from Lauer *et al*. [12]), with shape parameter 6 and scale parameter 0.88.

The distribution of infectiousness relative to symptom onset was based on a Gamma distribution with shape parameter 97.2 and scale parameter 0.2689, shifted by 25.6 days [13, 14]. It had a fixed four day pre-symptomatic phase, followed by a fixed ten day symptomatic phase. This gave a total of 14 days of infectivity and a minimum 15 day infection duration. The infectiousness temporal profile weighted the transmission risk (see Section 2.2.4) across the duration of the infectious period (for the full temporal profile, see Table 1). Following completion of the infectious period, the individual entered the recovered state. We provide a schematic representation of the epidemiological model in Fig. S1.

#### 2.2.2 Probability of asymptomatic infection and relative infectiousness

Infected individuals could be either asymptomatic throughout infection or symptomatic (*i*.*e*. develop symptoms following the latent period), according to a specified asymptomatic probability. Partitioning those infectious by symptom status allowed for the inclusion of a lower level of transmission believed to be associated with asymptomatic infection, and also a differential propensity to be identified as an infected case and self-isolate (see Section 2.3).

We allowed for the probability of asymptomatic infection to be age dependent (split into children and adolescents, 0-19yrs, and adults, 20+yrs). In each simulation run we sampled an asymptomatic infection probability for the 0-19yrs age group from a Uniform(0.2,0.35) and an asymptomatic infection probability for the 20+yrs age group from a Uniform(0.05,0.2) distribution; the width of these distributions were comparable to the 95% confidence intervals reported in the meta-analysis by Buitrago-Garcia *et al*. [15] for the estimated proportion of SARS-CoV-2 infections that were asymptomatic in studies of hospitalised children (95% confidence interval: 22%–32%) and hospitalised adults (95% confidence interval: 6%–19%), respectively (note that the meta-analysis by Buitrago-Garcia *et al*. [15] also suggested that the proportion of asymptomatic SARS-CoV-2 infection estimated in studies of hospitalised patients was similar to that in all other settings). The sampled asymptomatic probabilities remained fixed throughout that particular simulation run.

There was limited data available to provide a robust quantitative estimate of the relative infectiousness of asymptomatic and symptomatic individuals, with some support for the reduced infectiousness of asymptomatic individuals [15, 16]. Therefore, we assumed that asymptomatic individuals had a lower risk of transmitting infection compared to symptomatic individuals. To reflect the uncertainty in this area, for each simulation we sampled the relative infectiousness of asymptomatic individuals compared to symptomatic individuals from a Uniform(0.3, 0.7) distribution. This was sampled independently to the asymptomatic probability. The sampled value was applied as a scaling on transmission risk, applied evenly throughout the duration of infectiousness (*i*.*e*. with no time dependence, see Section 2.2.5).

#### 2.2.3 Susceptibility

Based on available estimates at the time, we set the relative susceptibility to SARS-CoV-2 infection for the child and adolescent age group (0-19yrs) to be approximately half that of people aged 20+yrs [17]. Explicitly, for each simulation we sampled the relative susceptibility of the child and adolescent age group (0-19yrs) from a Uniform(0.4, 0.6) distribution.

#### 2.2.4 Transmission risk

We used a data-driven approach to obtain the relative risk of transmission within households of differing composition.

To obtain the transmission risk from an individual, we modulated the infectivity profile using estimates of adjusted household secondary attack rates from a UK based surveillance study [18]. We attributed a household secondary attack rate to each person based on the size of their household. We sampled the attack rates for each individual from a normal distribution with mean dependent on the household size [18]: 0.48 for a household size of two, 0.40 for three, 0.33 for four, and 0.22 for five or more. The standard deviation of the normal distribution for households of size two or three was 0.06, and for households of four or more was 0.05. It should be noted that these estimates were made using a sample of 379 confirmed COVID-19 cases, meaning the robustness of the central estimates could be low. As such, we sampled from the previously mentioned normal distributions to ensure this uncertainty was captured.

On a day where households visited one another we assumed they would be gathered for the entire day. The overall transmission risk on a given day was dictated either by the total number of individuals in the household if there was no mixing with other households, or if multiple households were meeting as part of a support bubble or a Christmas bubble then the transmission risk was governed by the total number of individuals in the gathering. For example, if in a Christmas bubble there were three households of sizes 2, 1 and 1, then we assumed the transmission risk to be like a household of size 4.

#### 2.2.5 Probability of transmission per contact

We outline here how we used the household transmission risk, relative infectiousness of the infector, relative susceptibility of the infectee and the infectiousness temporal profile to compute the probability of transmission across an infectious-susceptible contact pair.

For an infectious individual *j* on day *t* of their infectious state, the probability of transmission to each susceptible contact *k* in household bubble *h*, denoted *p*_*j,k,h*_(*t*), was given by the product of four components:

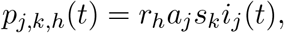

where *r*_*h*_ is the sampled household attack rate in household *h, a*_*j*_ the relative infectiousness of individual *j* (taking either value 1 for cases that were symptomatic during the infection episode, or the sampled value for relative infectiousness of asymptomatics compared to symptomatics otherwise), *s*_*k*_ the relative susceptibility of individual *k* and *i*_*j*_(*t*) the value of the infectiousness temporal profile on day *t* for individual *j* (Table 1).

### 2.3 Testing, contact tracing and isolation

Other measures in active use during December 2020 included symptomatic testing, contact tracing and isolation [19, 20]. We next expand on our implementation of the testing, contact tracing and isolation components within our household model. Associated parameters are listed in Table 2.

**Table 2:**
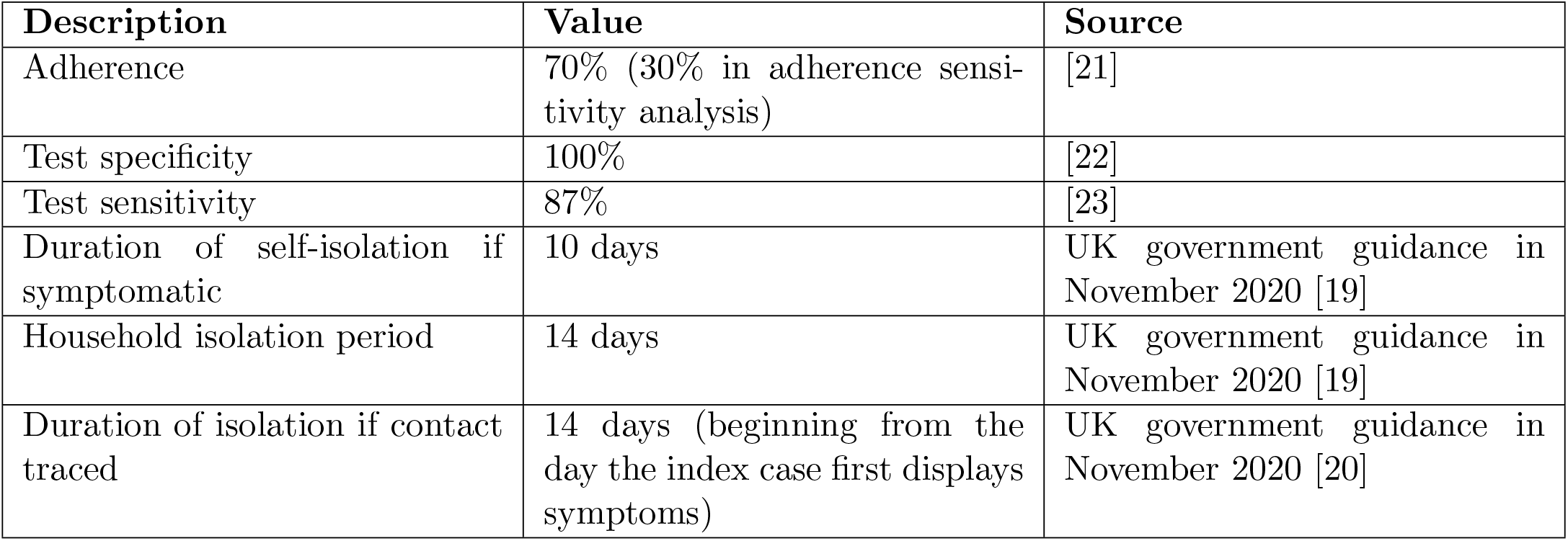
Description of testing and isolation related parameters.

| Description | Value | Source |
| --- | --- | --- |
| Adherence | 70% (30% in adherence sensitivity analysis) | [21] |
| Test specificity | 100% | [22] |
| Test sensitivity | 87% | [23] |
| Duration of self-isolation if symptomatic | 10 days | UK government guidance in November 2020 [19] |
| Household isolation period | 14 days | UK government guidance in November 2020 [19] |
| Duration of isolation if contact traced | 14 days (beginning from the day the index case first displays symptoms) | UK government guidance in November 2020 [20] |

#### 2.3.1 Adherence

We used an adherence parameter to capture the proportion of individuals that followed the recommended guidance. In our simulations all individuals within the same household (or extended household) had the same adherence status, representing whether an individual would both adhere to isolation guidance and engage with test and trace. We did not allow partial adherence to one measure and not the other. Adherence to isolation encompassed isolation for any reason: presenting with symptoms themselves, being in the same household as someone displaying symptoms, or being identified as a close contact of an infected individual via contact tracing.

We assumed a constant 70% adherence to isolation and test-and-trace measures, sampled using a Bernoulli trial. Though we acknowledge a subjective nature to this assumption, indicators from the Office for National Statistics (ONS) Opinions and Lifestyle Survey covering the period 05-08 November 2020 reported around 7 in 10 adults “finding it very easy or easy to follow the current lockdown measures where they live” [21]. To garner insights into the sensitivity to the adherence parameter we also performed simulations with a lower adherence, catering for a situation where the majority of the population were not partaking in other NPIs (see Section 2.5.4).

#### 2.3.2 Testing

We assumed an adherent individual household member took a PCR test if they displayed symptoms. We also assumed individuals would isolate between taking a PCR test and receiving the result. To enable a clearer assessment of the potential increase in infection resulting from the size and duration of Christmas bubbles, rather than possible confounding feedbacks that may be induced in the infection dynamics by lags in test result reporting, we assumed test results were delivered on the same day as symptom onset.

We set PCR test specificity at 100%, in line with data from the ONS COVID-19 Infection survey suggesting the false-positive rate to be very low, under 0.005% [22]. PCR test sensitivity was a fixed constant of 87%, based on the reported clinical sensitivity in symptomatic individuals [23], which also lay within the estimated range of 85% and 98% sensitivity reported across other studies [22]. These statistics agreed with the available estimates in the UK as of November 2020. That being said, at a similar time we were seeing the first reports in the literature that provided information on the sensitivity of PCR testing for SARS-CoV-2 infections dependent upon time since infection (studies such as Hellewell *et al*. [24], Wikramaratna *et al*. [25], for example). When carrying out this analysis we had yet to incorporate these time-dependent sensitivity profiles into the model.

In instances where the symptomatic household member returned a negative test result, they remained in self-isolation if they had independently been identified via contact tracing as a contact of a known infected; otherwise, the symptomatic individual left self-isolation. This matches government guidance on contact tracing at the time (see Section 2.3.3).

Household members were also released from isolation upon a negative result, as long as no other symptomatic cases (that were confirmed positive) were present in the household and they had not been independently identified as requiring to self-isolate through contact tracing.

#### 2.3.3 Contact tracing

Contacts of individuals returning a positive test result were retrieved as far back as two days prior to the emergence of symptoms for the index case. Contacts of a confirmed case were required to spend up to 14 days in self-isolation [20], matching the UK government guidance in November 2020. We set the isolation period to elapse 14 days after the index case became symptomatic.

#### 2.3.4 Isolation

Within an adherent household, symptomatic infected individuals isolated for 10 days upon symptom onset. All other individuals in the extended household were set to self-isolate for 14 days from their infected household members data of symptom onset [19]. Individuals in isolation did not participate in planned household visits due to occur during the five-day bubble window.

### 2.4 Christmas bubble scenarios

We evaluated five Christmas bubble scenarios, which could be in use during 23-27 December 2020. The five scenarios, ranging from least to greatest amount of unique additional contacts, were:

**Scenario A - No change to household bubble guidance:** Only support bubbles (extended households) permitted to meet each day.

**Scenario B - Short duration fixed exclusive bubbles:** The same three households meet each day between 25-26 December 2020, with no overlapping between bubbles.

**Scenario C - Fixed exclusive bubbles:** The same three households meet each day between 23-27 December 2020, with no overlapping between bubbles. This scenario most closely corresponds to the original Christmas bubble plan for England.

**Scenario D - Fixed non-exclusive bubbles:** Each household meets two other households each day between 23-27 December 2020. Bubbles may overlap.

**Scenario E - Daily change in household triplets:** Each day, all households are assigned to a household triplet. The household triplets change each day between 23-27 December 2020.

In all scenarios we assumed extended households (households part of a support bubble) mixed every day during 23-27 December 2020. Furthermore, in Scenarios B-E existing support bubbles counted as one household towards the three household limit, concurring with the original Christmas bubbles guidance [3]. For a visual illustration of the different scenarios, see Fig. 1.

**Fig. 1:**
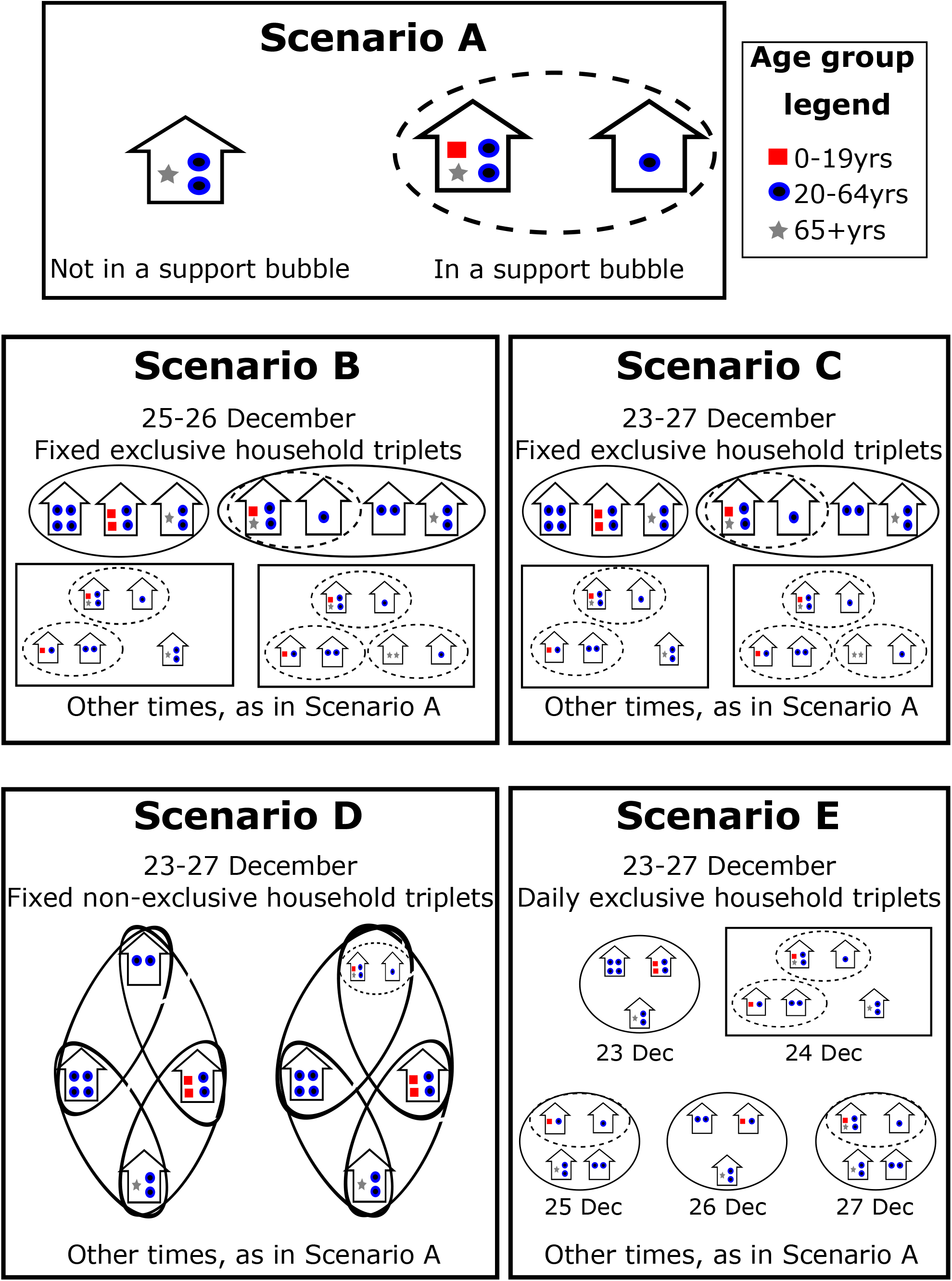
Illustration of the modelled Christmas bubble scenarios. We depict our five modelled scenarios. Scenario A: No change (support bubbles only); Scenario B: Short duration fixed exclusive bubbles, meet 25 and 26 December only; Scenario C: Fixed exclusive bubbles, meet every day 23-27 December; Scenario D: Fixed non-exclusive bubbles, meet every day 23-27 December; Scenario E: New household triplets meet each day between 23-27 December. Households contained within dashed ellipses signify an existing support bubble. Households within solid ellipses or rectangles are households contacted as part of a Christmas bubble. In Scenarios B-E, existing support bubbles counted as one household towards the three household limit. We use markers to denote individuals and different marker types signify individuals in different age groups; 0-19yrs (red squares); 20-64yrs (blue circles); 65+yrs (grey stars).

### 2.5 Simulation overview

#### 2.5.1 Model initialisation

For each scenario we performed 100 model runs on a system containing 100,000 households, with an approximate overall population of 310,000 people. These quantities were pragmatic choices, balancing between generating sufficient realisations spanning the plausible range of parameter values and having a large overall population size for which epidemic simulations could be completed in a confined time frame.

At the outset of each simulation we assigned household bubbles in a two-step process. Firstly, for each single-occupancy household we checked whether they would form a support bubble with another household (see Section 2.1). Secondly, for Scenarios B-E, we created Christmas bubbles and visitation schedules according to the specifics of each scenario. The second step of the household bubble allocation was not applicable for Scenario A.

We then proceeded to initialise the epidemiological state of each individual. Accordingly, we assigned to each individual one of the following possible disease statuses; susceptible, latently infected, asymptomatically infected, presymptomatic infectious, recovered. There were intentionally no individuals initialised as symptomatically infected at the start of the simulated time horizon, meaning no households began in isolation.

The probability of being attributed a given disease state was age-dependent, with age-stratified estimates for our three age groups (0-19yrs, 20-64yrs, 65+yrs) informed by inferred values from the Warwick SARS-CoV-2 compartmental transmission model in late November 2020 [26]. We set the status of individuals in each household at random, with acknowledgement that in truth disease statuses within a household may be correlated due to within household transmission (remarked upon further in the Discussion). We provide initial condition values in Table 3.

**Table 3:** Percentage of each age group initialised in each infection status.

|  | Age (years) |  |  |
| --- | --- | --- | --- |
|  | 0-19 | 20-64 | 65+ |
| **Susceptible** | 73% | 74% | 84.5% |
| **Latent infected** | 1% | 0.5% | 0.25% |
| **Asymptomatic infected** | 0.3% | 0.1% | 0.05% |
| **Presymptomatic infected** | 0.7% | 0.4% | 0.2% |
| **Recovered** | 25% | 25% | 15% |

#### 2.5.2 Main analysis

For our main analysis we began simulations on 23 December 2020 and and ran each simulation for 15 days. During 23-27 December 2020 contacts were made according to the Christmas bubble scenario being studied. After 27 December 2020, we assumed extended households continued to interact each day for ten further days.

The described setup corresponded to a population of households that had no symptomatic infection present, and therefore no households being in isolation, at the beginning of the five-day Christmas bubble window on 23 December 2020.

#### 2.5.3 Sensitivity analysis: Alternative time horizon

As a sensitivity analysis, we considered an alternative set of circumstances where it was possible for households to be in isolation as the Christmas bubble period began.

In this case we began the simulations ten days prior to 23 December 2020 (*i*.*e*. simulations began on 13 December 2020), with the same initial conditions as used for the main analysis. On each day during 13-22 December 2020 extended households would meet each day. For 23-27 December 2020 we ran the previously described set of five Christmas bubble scenarios. As before, after 27 December 2020 we assumed extended households continued to interact each day for ten further days.

The intent of this setup was to more closely reflect the likely epidemiological impacts averaged over the whole general population, rather than the epidemiological impacts amongst solely households that had no symptomatic infection present and requirement to isolate upon the temporary relaxation of social restrictions.

#### 2.5.4 Sensitivity analysis: Adherence to test, trace and isolation measures

Lastly, for both the main and alternative analyses time horizons we investigated epidemiological outcomes given a lower adherence to isolation and test-and-trace measures of 30% (rather than 70%).

#### 2.5.5 Epidemiological outcomes

Our assessment of the Christmas bubble and visitation scenarios primarily considered a pair of incidence and cumulative infection statistics. We grouped these epidemiological measures into: (i) Daily incidence each day between 23-27 December 2020; (ii) Cumulative infections over the entire modelled time horizon. For these quantities we were also interested in the share attributable to each age group.

#### 2.5.6 Software and code

We performed the model simulations in Julia v1.5 and generated the figures in Matlab R2022a. The code repository for the study is available at: https://github.com/EdMHill/SARS-CoV-2_Christmas_bubbles_2020.

## 3 Results

### 3.1 Longer and larger visits alters the age distribution of cases

We visualise the daily incidences from 23 December 2020 (Fig. 2(a)) to 27 December 2020 (Fig. 2(e)), for all five Christmas bubble scenarios ordered from the lowest level of unique contacts (Scenario A, darkest colour) to the highest level (Scenario E, lightest colour), and sorted by age group (blue, left across to purple, right). We particularly note the dynamics exhibited by Scenario B that allowed exclusive bubbles of three households on 25 and 26 December 2020 only.

**Fig. 2:**
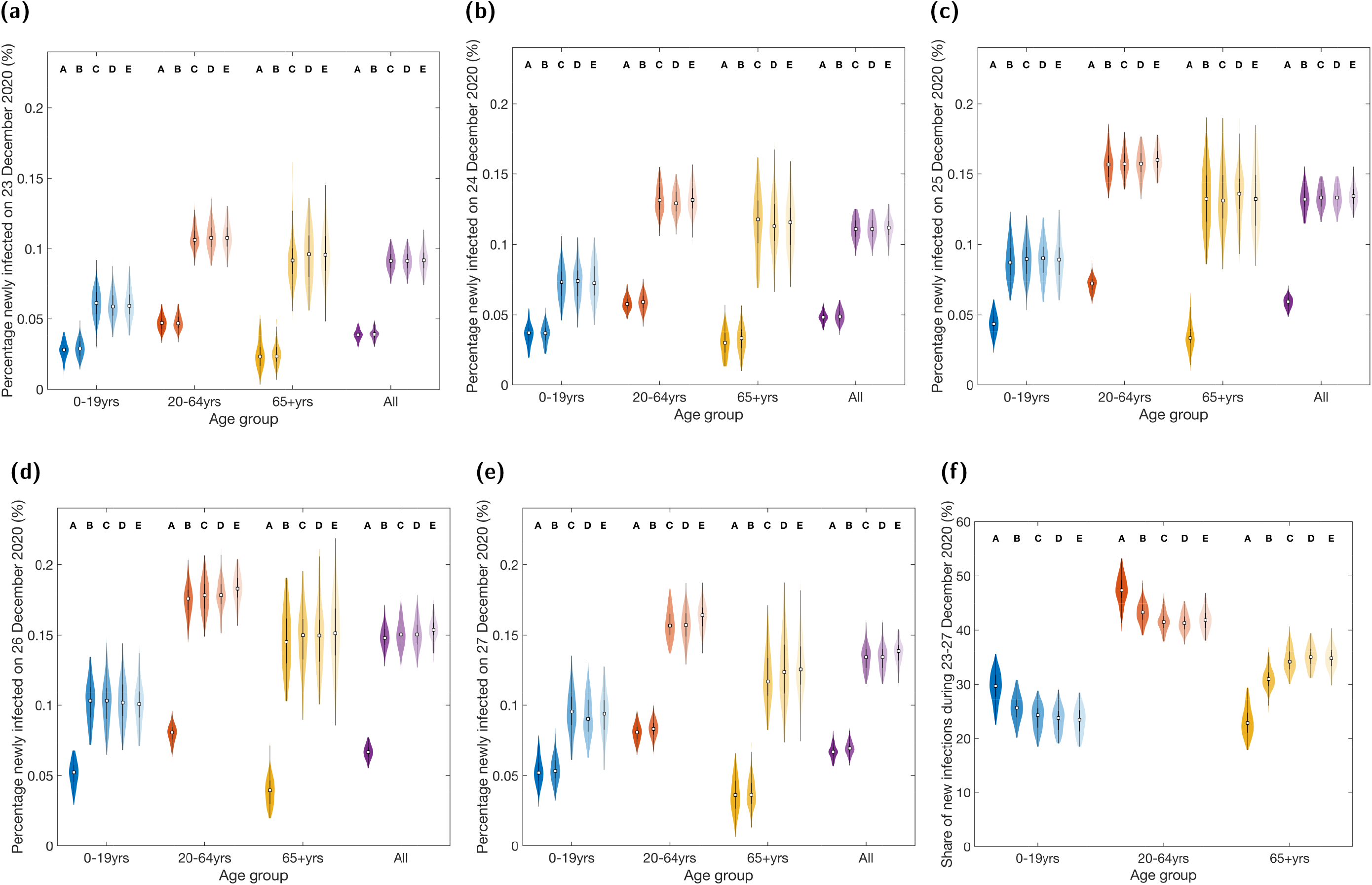
Distributions for the daily incidence and age-stratification of new infections between 23-27 December 2020 under each Christmas bubble scenario. Estimates produced from 100 realisations per scenario for daily incidence on: **(a)** 23 December 2020; **(b)** 24 December 2020; **(c)** 25 December 2020; **(d)** 26 December 2020; **(e)** 27 December 2020. Panel **(f)** displays the share of new infections between 23-27 December 2020 attributable to each of the three age groups. In each violin plot the white squares represent the medians and solid black lines correspond to the interquartile range. See Table S1 for median values and 95% prediction intervals. Labels A-E correspond to the distributions associated with Scenarios A-E. The intensity of shading of the violin plots also distinguishes between the scenarios; darkest for Scenario A to lightest for Scenario E. Scenario A: No change (support bubbles only); Scenario B: Short duration fixed exclusive bubbles, meet 25 and 26 December only; Scenario C: Fixed exclusive bubbles, meet every day 23-27 December; Scenario D: Fixed non-exclusive bubbles, meet every day 23-27 December; Scenario E: New household triplets meet each day between 23-27 December. Relative to Scenario A, the increase in incidence for Scenarios C-E was greatest for the 65+yrs age group.

For 23 and 24 December 2020, Scenario B had the same rules as Scenario A, and thus similar daily incidence distributions (Figs. 2(a) and 2(b), Table S1). For the two days where Christmas bubbles were implemented (25 and 26 December 2020), the daily incidence distributions were similar to Scenarios C-E, where households were allowed to meet two other households on all five days between 23-27 December 2020 (Figs. 2(c) and 2(d), Table S1). On 27 December 2020, the incidence returned to levels comparable to Scenario A, though median estimates for Scenario B were now slightly larger (Fig. 2(e), Table S1); the overall estimates for all ages in the population were 0.067% (95% PI:0.058%- 0.079%) for Scenario A, compared to 0.069% (95% PI: 0.061%-0.080%) for Scenario B.

We next remark on an apparent age-dependence for the expected increase in incidence on days when Christmas bubbles were permitted to meet. Compared to maintaining support bubbles only (Scenario A), on days Christmas bubbles could meet we found the incidence approximately trebled for those aged 65+yrs. In contrast, for the 20-64yrs age group incidence roughly doubled, whereas the relative change was smallest for those aged 0-19yrs (Figs. 2(a) to 2(e)).

We observed a shift towards a greater proportion of infections occurring in the 65+yrs age group (Fig. 2(f)) as a result of the use of exclusive bubbles of three households for a short duration (Scenario B). Under Scenario A, the percentage of new infections during this five day window estimated to occur in those aged 65+yrs was 23% (95% PI: 19%-29%). Under Scenario B, the analogous median and uncertainty estimates were 31% (95% PI: 27%-35%). On the other hand, the 0-19yrs and 20-64yrs age groups had similar declines in contribution to new infections during that time period (when comparing Scenario B to Scenario A).

With an increase in duration of interaction (Scenarios C and D) or the number of contacts (Scenario E), we observed a larger shift in the age distribution of new infections that occurred during the period 23-27 December 2020. Specifically, there was an appreciable rise of roughly 10% in the proportion of cases attributed to those aged 65 and above, with a corresponding decrease in the region of 5% of the share of new infections attributable to those aged 0-19yrs and 20-64yrs (Fig. 2(f)).

### 3.2 Implications of Christmas bubbles on post-Christmas SARS-CoV-2 prevalence

All Christmas bubble scenarios that allowed for mixing beyond extended households resulted in an increase in the amount of new infections over the whole 15-day time horizon, 23 December 2020 to 06 January 2021 (Fig. 3). Notably, the total number of days spent mixing within that period could impact post-Christmas prevalence. Compared to having no additional household bubbling (Scenario A), for which cumulative infections for all ages reached 0.60% (95% PI: 0.53%-0.64%), the rises in cumulative infections were most stark for household visit schedules where three households could meet each day for the full five days (Scenarios C-E).

**Fig. 3:**
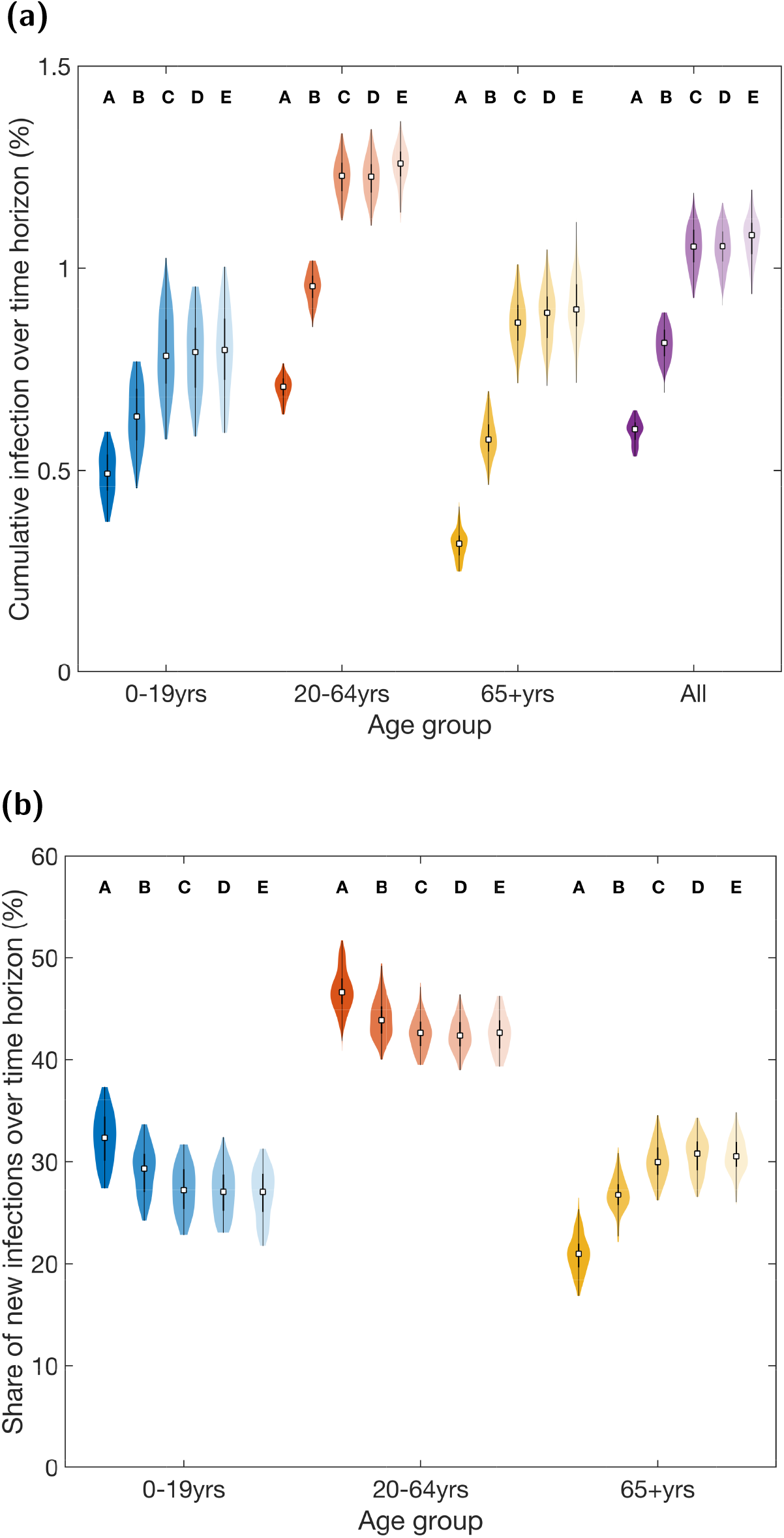
Cumulative infection distributions for the entire 15 day time horizon, 23 December 2020 to 06 January 2021, under each Christmas bubble scenario. Estimates produced from 100 realisations per scenario. **(a)** The percentage of each age group newly infected during the time horizon. **(b)** The percentage of infections during the time horizon attributed to each age group. In each violin plot the white squares represent the medians and solid black lines correspond to the interquartile range. See Table S2 for median values and 95% prediction intervals. Labels A-E correspond to the distributions associated with Scenarios A-E. The intensity of shading of the violin plots also distinguishes between the scenarios; darkest for Scenario A to lightest for Scenario E. Scenario A: No change (support bubbles only); Scenario B: Short duration fixed exclusive bubbles, meet 25 and 26 December only; Scenario C: Fixed exclusive bubbles, meet every day 23-27 December; Scenario D: Fixed non-exclusive bubbles, meet every day 23-27 December; Scenario E: New household triplets meet each day between 23-27 December. All scenarios beyond extended household bubbles resulted in an increase in the amount of new infections amongst the population over the considered time horizon.

In detail, we found modelled cumulative infections across all ages of: exclusive bubbles (Scenario C) - 1.05% (95% PI: 0.95%-1.15%); non-exclusive fixed bubbles (Scenario D) - 1.05% (95% PI: 0.95%- 1.15%); daily change in household triplets (Scenario E) - 1.08% (95% PI: 0.94%-1.18%). We remark that although Scenario E, with new household triplets meeting each day of 23-27 December 2020, would result in a greater amount of distinct person-to-person contacts being made compared to having fixed bubbles comprised of three households (Scenarios C and D), the modelled outcomes across these three scenarios were similar. On the other hand, restricting the duration of exclusive bubbles to 25-26 December 2020 (Scenario B) reduced the relative increase in cumulative infections over the 15-day time horizon by approximately half (Fig. 3, Table S1).

Stratifying by age, the greatest relative increase in cumulative infection for Scenario E versus Scenario A was amongst the eldest age group (Scenario A - 0.32% (95% PI: 0.26%-0.39%), Scenario E - 0.90% (95% PI: 0.75%-1.06%), Fig. 3). The 0-19yrs child and adolescent age group had the lowest relative increase (Scenario A - 0.49% (95% PI: 0.38%-0.58%), Scenario E - 0.80% (95% PI: 0.60%-0.98%)).

### 3.3 Sensitivity analysis: Alternative time horizon

We next inspected our simulations that began on 13 December 2020, which included households with initially symptomatic individuals and those already in isolation on 23 December 2020, the start of the (up to) five-day temporary easement of social restrictions. One may consider such a setting to be more analogous to the likely state of the general population as Christmas bubbles began.

We observed qualitatively similar findings to the main analysis when inspecting epidemiological outcomes both constrained to the 23-27 December 2020 period (Fig. S2, Table S3) and over the entire time horizon through to 06 January 2021 (Fig. S3, Table S4). Shorter duration exclusive bubbles (Scenario B) roughly halved the additional infections accumulated compared to Scenarios C-E where meeting with two other households was allowed for all five days during 23-27 December 2020 (Fig. S3(a)). Relative to maintaining the status quo of support bubbles (Scenario A), allowing in the period 23-27 December 2020 fixed exclusive bubbles of three households to meet all five days (Scenario C), fixed non-exclusive bubbles of three households to meet all five days (Scenario D), or on each day different household triplets to meet (Scenario E) resulted in a higher proportion of cases occurring in the 65+yes age class (Figs. S2(f)&S3(b)). These three bubbling scenarios also caused similar levels of additional infections (Figs. S2&S3).

There were four quantitative differences between the outcomes for our alternative analysis compared to the main analysis (see Tables S3-S4 for summary statistics). First, in the alternative analysis the daily incidence during 23-27 December 2020 for each Christmas bubble scenario and age group combination was of the order of five-fold less compared to the corresponding scenario in the main analysis. Second, independent of the Christmas bubble strategy in use, the 0-19yrs age group had a higher relative share of new infections. Third, the magnitude of the relative increase in cumulative infections compared to there being no temporary easement in social restrictions (Scenario A) was less for household visit schedules that resulted in interactions of longer duration (Scenarios C and D) or a greater amount of contacts (Scenario E). Finally, there was greater variability in all predicted distributions (Figs. S2&S3).

### 3.4 Sensitivity analysis: Adherence to test, trace and isolation measures

The last piece of analysis investigated sensitivity of the epidemiological outcomes to the adherence parameter. We considered circumstances where the majority of the population were not adherent to isolation and test-and-trace measures, setting the adherence probability to 30%.

As anticipated, irrespective of the Christmas bubble scenario and age group, lower adherence led to higher incidence and cumulative infections. However, we still found qualitatively similar findings regarding age distributions, chiefly a marked uplift in the proportion of infections occurring in those aged 65 and above (Figs. S4-S7, Tables S5-S8).

## 4 Discussion

In this study, we have developed a stochastic individual-based model to analyse the spread of SARS-CoV-2 in a population experiencing a temporary easement in social restrictions, permitting the creation of bubbles of three households. We have investigated the impact of different bubble strategies, akin to the ‘Christmas bubbles’ proposed for use in England between 23-27 December 2020, upon SARS-CoV-2 incidence and prevalence amongst a synthetic population of households.

We found incidence and cumulative infection metrics were similar for bubble policies allowing either longer duration meetings of fixed household bubbles or multiple one-day gatherings of unique household triplets. Further, it was likely that visiting family and friends over the holiday period for two days would be less risky than spending the entire time together; shorter and smaller gatherings reduced the relative amount of infection. We also observed the increases in infection from greater amounts of social mixing disproportionately impacted the eldest. This is of particular concern for SARS-CoV-2 infections due to the worse clinical outcomes associated with increasing age, with higher hospitalisation and mortality rates of COVID-19 disease observed in older age groups ages [2, 27]. These conclusions were maintained for a population of households where households could begin the Christmas bubble time period in isolation, thus fragmenting the potential maximum connectivity that usage of Christmas bubbles could bring, and in a setting where the majority of the population would not be adherent to test, trace and isolate guidance.

Our findings have since been corroborated by subsequent additions to the literature on the topic of quantifying the effect of household bubbles on the transmission of SARS-CoV-2, with a variety of modelling approaches represented [28, 29]. In detail, Danon *et al*. [28] used the configuration model to link households and generate a network description of households in the UK. They found that whilst the creation of support bubbles between a single-person household and another household of any size had a small impact on transmission, scenarios where all households form a bubble would be highly likely to cause extensive transmission. Later work from Hilton *et al*. [29] provided an infectious disease modelling framework formulated in terms of tractable systems of ordinary differential equations that includes explicit representation of age/risk structure and household structure. Our results are in agreement with their findings on the usage of short-term social bubbles, which suggested that short-term relaxation in mixing restrictions would have a small but non-negligible impact on the epidemic dynamics, with larger temporary bubbles and longer mixing periods associated with higher prevalence. On this basis, we would anticipate that if we had a finer-grain age stratification in our model than the three age groups used in this study (0-19yrs, 20-64yrs, 65+yrs), we would obtain qualitatively similar outcomes.

The model framework described here is reasonably parsimonious compared to fine-detailed household models [29–32]. With regards to informing policy in response to a global public health emergency such as a pandemic, parsimonious models that can be promptly set up and that are able to generate results in extremely short time-scales can be helpful tools for providing high-level insights, on the basis that a reasonable but imperfect answer is better than none [33].

Furthermore, there is scope for the model to be adapted and deployed to investigate similar questions given the contemporary SARS-CoV-2 epidemiological context, though the model framework would first require modification. Notable modifications that would be needed include accounting for vaccination status, waning of immunity and varying transmissibility to reflect the changes in dominant variants in community circulation (with this study having only considered a single variant). Another data-driven update would be to the infectivity profile estimates, with profiles now available that are UK specific [34] and variant specific [35].

The model also has ongoing prospective use in situations where substantial mixing between household groups is expected over a short duration of time, in relation to between-household contact levels before and after the given mixing period. Such situations are of interest as the period of increased mixing between household groups can result in a new transmission network as additional connections are made. In circumstances where the network then returns to normal work and school connections afterwards, infection can potentially be introduced into new groups. These effects, however, would be uncertain and remain challenging to resolve quantitatively in real-time.

Where possible, we have taken a data-driven approach to parameterise the system and instruct heterogeneities we expect to be present, such as in age-dependent susceptibility and household composition by age group. Nevertheless, this work has made simplifying assumptions and our results therefore have limitations.

We only considered infection resulting from person-to-person interactions due to household mixing. We did not consider transmission arising from other settings, with it also possible that the formation and ceasing of bubbles may have additional effects on transmission. As an example, infections could be seeded in households due to external contacts arising from the workplace (although there would typically be fewer people attending the workplace during the Christmas and New Year period). Such connectivity bridging households would naturally result in a greater number of contacts being made overall and likely hasten the spread of infection. Perhaps counterintuitively, it could also result in households entering isolation earlier and a more immediate curbing of the outbreak. These nonlinearities exhibited between isolation levels and infection burden merit further exploration.

Our findings may be sensitive to alternative epidemiological model structures and intervention assumptions. Of particular note is adherence to isolation and test-and-trace measures. By November 2020 there was growing evidence for nationally applied NPIs (such as social distancing, self-isolation upon symptom onset and household quarantine) having helped reduce the spread of SARS-CoV-2 [36–38]. Though we provide a sensitivity analysis to the adherence parameter (of 30% compared to the value of 70% used in the main analysis), conducting the evaluation of different Christmas bubble options within the time compressed needs of decision making in the policy arena was the priority. The true adherence of the population, and how this could change over time, warrants continued appraisal.

Our assumptions regarding bubble formation were also a simplified representation of the real-world social system. First, we assumed all households would join a Christmas bubble, which would not necessarily be the case. Second, we did not include the possibility of pre-existing household units splitting apart to join their own independent bubbles over the course of the five day window. Third, we did not investigate clustered initial infections (which were instead seeded randomly in the population). Finally, we did not consider childcare bubbles [39]. Childcare bubbles provided informal childcare (informal in the sense the childcare was unregistered) for anyone under 14 from friends and/or family from one other household. Stipulations included only having one childcare bubble with one other household at a time, meaning no household could be part of more than one childcare bubble. The inclusion of these other types of allowed household bubbles would alter the plausible combinations of households that may form a Christmas bubble. Our findings should be interpreted with due consideration of the pragmatic decisions made to ensure analyses could be delivered in a timely manner.

Finally, the original motivation for this scenario modelling study was to help inform decision-making and our understanding of the SARS-CoV-2 epidemic in England during December 2020. Another route of further work is validation of the model with the subsequent empirical data, though there are challenges to be surmounted to enable a suitable ‘baseline model’ to be selected (as there were regional variations in the social restrictions applied in England during the Christmas period in 2020 [6]) and remove artefacts from the empirical data caused by multiple bank holidays over the festive period disrupting surveillance data streams. Additionally, it should be recognised that comparisons between modelled scenarios and what actually happens following a decision are less informative about the scenario modelling itself than they are about the impacts of these changes and the important processes driving the epidemic [40].

Models of infectious disease transmission are able to provide quantitative evidence for the potential impact of different intervention options on the future course of an outbreak. This study demonstrates the potential uses of a stochastic individual-based model representing a synthetic population of households, which could be developed swiftly and deployed in situations necessitating assessment of the epidemiological impact of extending contacts beyond the immediate household. Our findings offer general insights on how short term changes to household bubbles might influence the infectious disease dynamics, particularly alterations to the age distributions of epidemiological outcomes of interest.

## Data Availability

All data utilised in this study are publicly available, with relevant references and data repositories stated within the main manuscript and Supporting Information. 
The code repository for the study is available at https://github.com/EdMHill/SARS-CoV-2_Christmas_bubbles_2020.
Archived code: https://doi.org/10.5281/zenodo.6794136

https://doi.org/10.5281/zenodo.6794136

## Acknowledgements

With thanks to Louise Dyson, Matt Keeling, Mike Tildesley and Robin Thompson for helpful discussions and comments on the manuscript.

## Author contributions

**Edward M. Hill:** Conceptualisation, Data curation, Formal analysis, Methodology, Software, Validation, Visualisation, Writing - Original Draft, Writing - Review & Editing.

## Data availability

All data utilised in this study are publicly available, with relevant references and data repositories stated within the main manuscript and Supporting Information.

## Code availability

The code repository for the study is available at https://github.com/EdMHill/SARS-CoV-2 Christmas bubbles 2020.

Archived code: https://doi.org/10.5281/zenodo.7134811

