## Supporting Information for "Modelling the epidemiological implications for SARS-CoV-2 of Christmas household bubbles in England"

Edward M. Hill[1,2*]

**1** The Zeeman Institute for Systems Biology & Infectious Disease Epidemiology Research, School of Life Sciences and Mathematics Institute, University of Warwick, Coventry, United Kingdom.
**2** Joint UNIversities Pandemic and Epidemiological Research, https://maths.org/juniper/.

### Table of Contents

**Additional figures** **2**

**Additional tables** **9**

### Additional figures

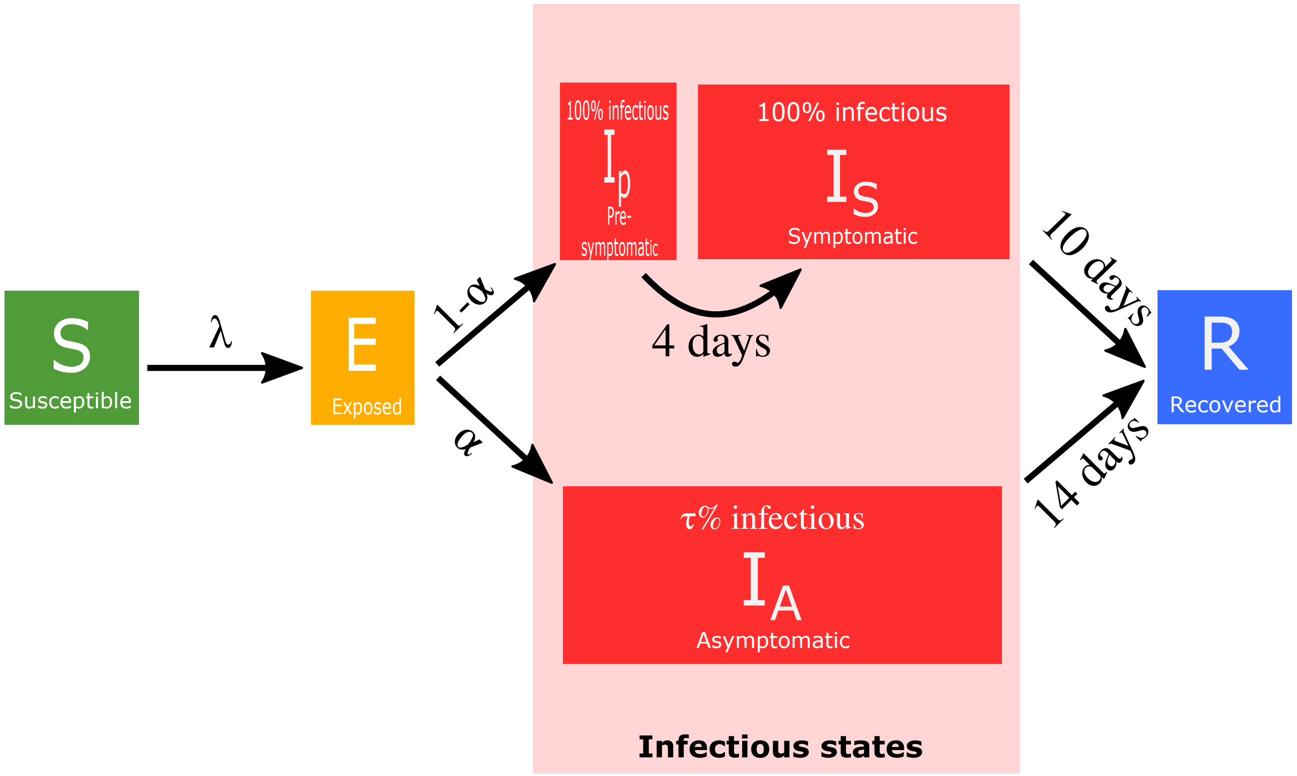


**Fig. S1: Epidemiological model disease states and transitions.** We stratified the population into susceptible ($S$), exposed ($E$, infected but not yet infectious), presymptomatic infectious ($I_p$), symptomatic infectious ($I_S$), asymptomatic infectious ($I_A$), and recovered ($R$) states. Solid lines denote transitions between disease states. A susceptible individual ($S$) became infected due to infectious pressure ($\lambda$) exerted by having contact with an infectious individuals, which can lead to onward transmission of the virus. Upon infection, individuals entered the exposed state ($E$). Upon leaving the exposed period, the individual entered the infectious state on one of two pathways: (i) with probability $\alpha \in [0.3, 0.7]$ they entered the asymptomatic state ($I_A$), with the parameter $\tau$ denoting the relative infectiousness of an asymptomatic versus a symptomatic case and presymptomatic cases that will become symptomatic. $\tau$ was dependent on age (0-19yrs: $\tau \in [0.20, 0.35]$; 20+yrs: $\tau \in [0.05, 0.20]$); (ii) with probability 1-$\alpha$ they entered the presymptomatic state ($I_p$), before becoming symptomatic ($I_S$) after four days. Upon resolution of infection individuals moved to the recovered ($R$) state. Infectious individuals left the infectious state after 14 days, meaning infectious cases that went on to become symptomatic remained in the symptomatic state for 10 days.

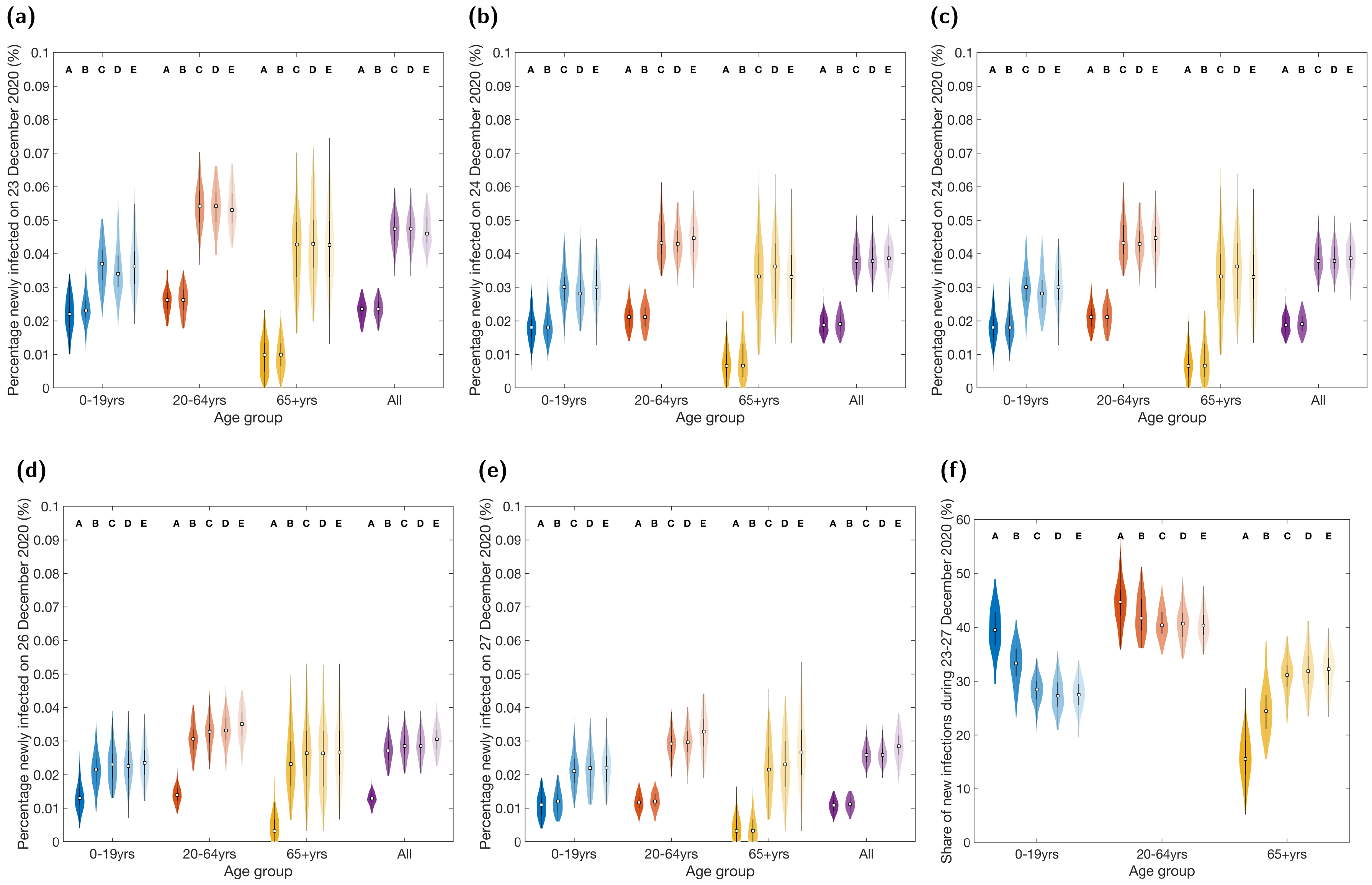


**Fig. S2: For the alternative analysis with simulations beginning on 13 December 2022, distributions for the daily incidence and age-stratification of new infections between 23-27 December 2020 under each Christmas bubble scenario.** Estimates produced from 100 realisations per scenario for daily incidence on: **(a)** 23 December 2020; **(b)** 24 December 2020; **(c)** 25 December 2020; **(d)** 26 December 2020; **(e)** 27 December 2020. Panel **(f)** displays the share of new infections between 23-27 December 2020 attributable to each of the three age groups. In each violin plot the white squares represent the medians and solid black lines correspond to the interquartile range. See Table S3 for median values and 95% prediction intervals. Labels A-E correspond to the distributions associated with Scenarios A-E. The intensity of shading of the violin plots also distinguishes between the scenarios; darkest for Scenario A to lightest for Scenario E. Scenario A: No change (support bubbles only); Scenario B: Short duration fixed exclusive bubbles, meet 25 and 26 December only; Scenario C: Fixed exclusive bubbles, meet every day 23-27 December; Scenario D: Fixed non-exclusive bubbles, meet every day 23-27 December; Scenario E: New household triplets meet each day between 23-27 December.

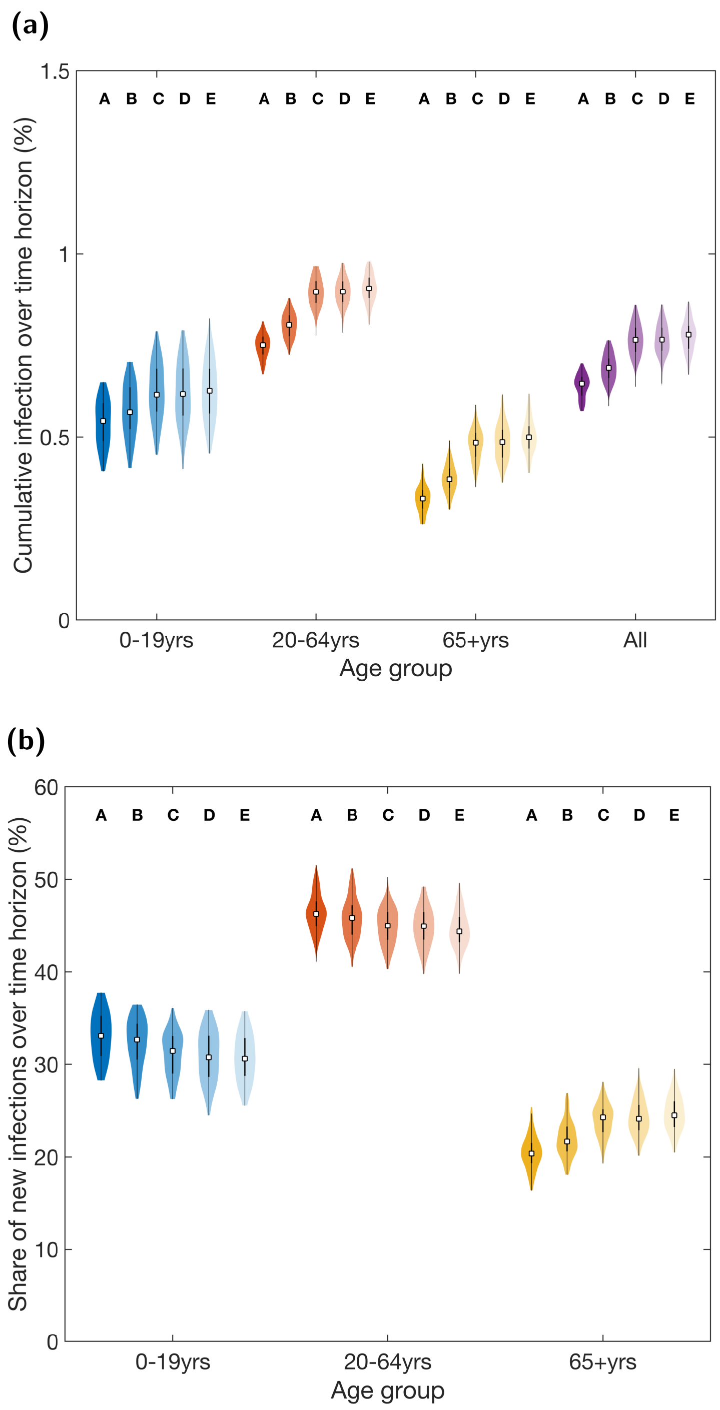


**Fig. S3: For the alternative analysis with simulations beginning on 13 December 2022, cumulative infection distributions for the entire 25 day time horizon, 13 December 2020 to 06 January 2021, under each Christmas bubble scenario.** Estimates produced from 100 realisations per scenario. **(a)** The percentage of each age group newly infected during the time horizon. **(b)** The percentage of infections during the time horizon attributed to each age group. In each violin plot the white squares represent the medians and solid black lines correspond to the interquartile range. See Table S3 for median values and 95% prediction intervals. Labels A-E correspond to the distributions associated with Scenarios A-E. The intensity of shading of the violin plots also distinguishes between the scenarios; darkest for Scenario A to lightest for Scenario E. Scenario A: No change (support bubbles only); Scenario B: Short duration fixed exclusive bubbles, meet 25 and 26 December only; Scenario C: Fixed exclusive bubbles, meet every day 23-27 December; Scenario D: Fixed non-exclusive bubbles, meet every day 23-27 December; Scenario E: New household triplets meet each day between 23-27 December.

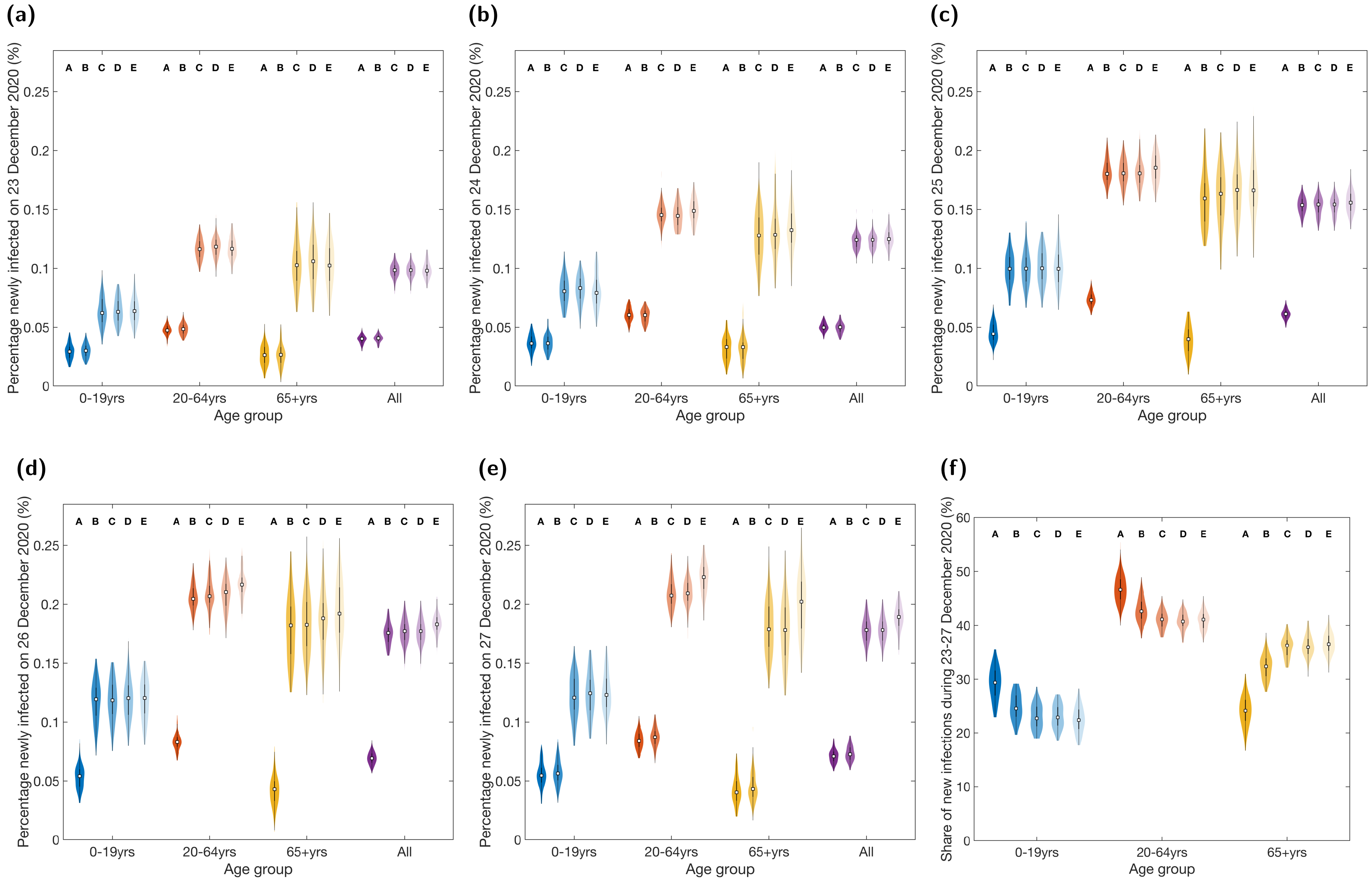


**Fig. S4: Distributions for the daily incidence and age-stratification of new infections between 23-27 December 2020 under each Christmas bubble scenario for the main analysis (with simulations beginning on 23 December 2022) combined with the lower adherence to testing, contact tracing and isolation measures assumption.** In these simulations, each household had a 30% probability of being adherent to the guidance on testing, contact tracing and isolation. Estimates produced from 100 realisations per scenario for daily incidence on: **(a)** 23 December 2020; **(b)** 24 December 2020; **(c)** 25 December 2020; **(d)** 26 December 2020; **(e)** 27 December 2020. Panel **(f)** displays the share of new infections between 23-27 December 2020 attributable to each of the three age groups. In each violin plot the white squares represent the medians and solid black lines correspond to the interquartile range. See Table S3 for median values and 95% prediction intervals. Labels A-E correspond to the distributions associated with Scenarios A-E. The intensity of shading of the violin plots also distinguishes between the scenarios; darkest for Scenario A to lightest for Scenario E. Scenario A: No change (support bubbles only); Scenario B: Short duration fixed exclusive bubbles, meet 25 and 26 December only; Scenario C: Fixed exclusive bubbles, meet every day 23-27 December; Scenario D: Fixed non-exclusive bubbles, meet every day 23-27 December; Scenario E: New household triplets meet each day between 23-27 December.

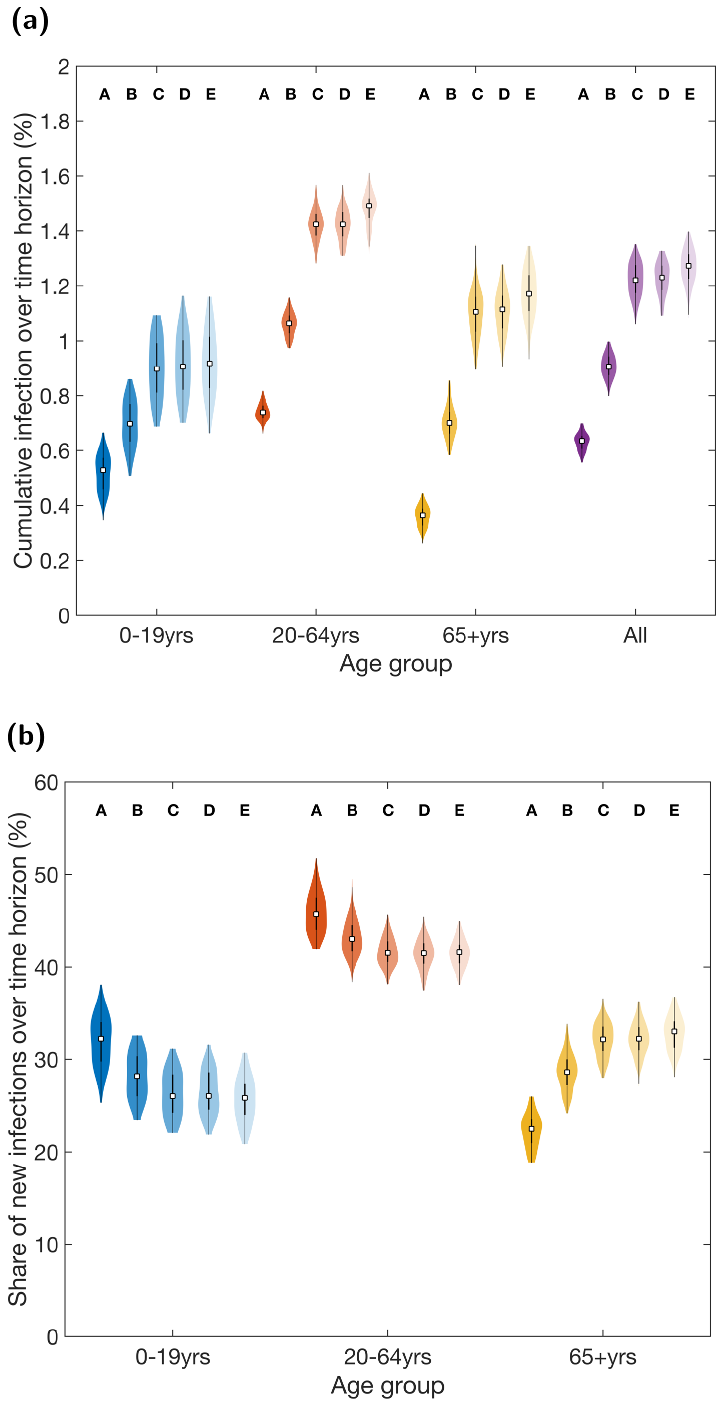


**Fig. S5: For the main analysis with simulations beginning on 23 December 2022 and lower adherence to testing, contact tracing and isolation measures, cumulative infection distributions for the entire 15 day time horizon, 23 December 2020 to 06 January 2021, under each Christmas bubble scenario.** In these simulations, each household had a 30% probability of being adherent to the guidance on testing, contact tracing and isolation. Estimates produced from 100 realisations per scenario. **(a)** The percentage of each age group newly infected during the time horizon. **(b)** The percentage of infections during the time horizon attributed to each age group. In each violin plot the white squares represent the medians and solid black lines correspond to the interquartile range. See Table S3 for median values and 95% prediction intervals. Labels A-E correspond to the distributions associated with Scenarios A-E. The intensity of shading of the violin plots also distinguishes between the scenarios; darkest for Scenario A to lightest for Scenario E. Scenario A: No change (support bubbles only); Scenario B: Short duration fixed exclusive bubbles, meet 25 and 26 December only; Scenario C: Fixed exclusive bubbles, meet every day 23-27 December; Scenario D: Fixed non-exclusive bubbles, meet every day 23-27 December; Scenario E: New household triplets meet each day between 23-27 December.

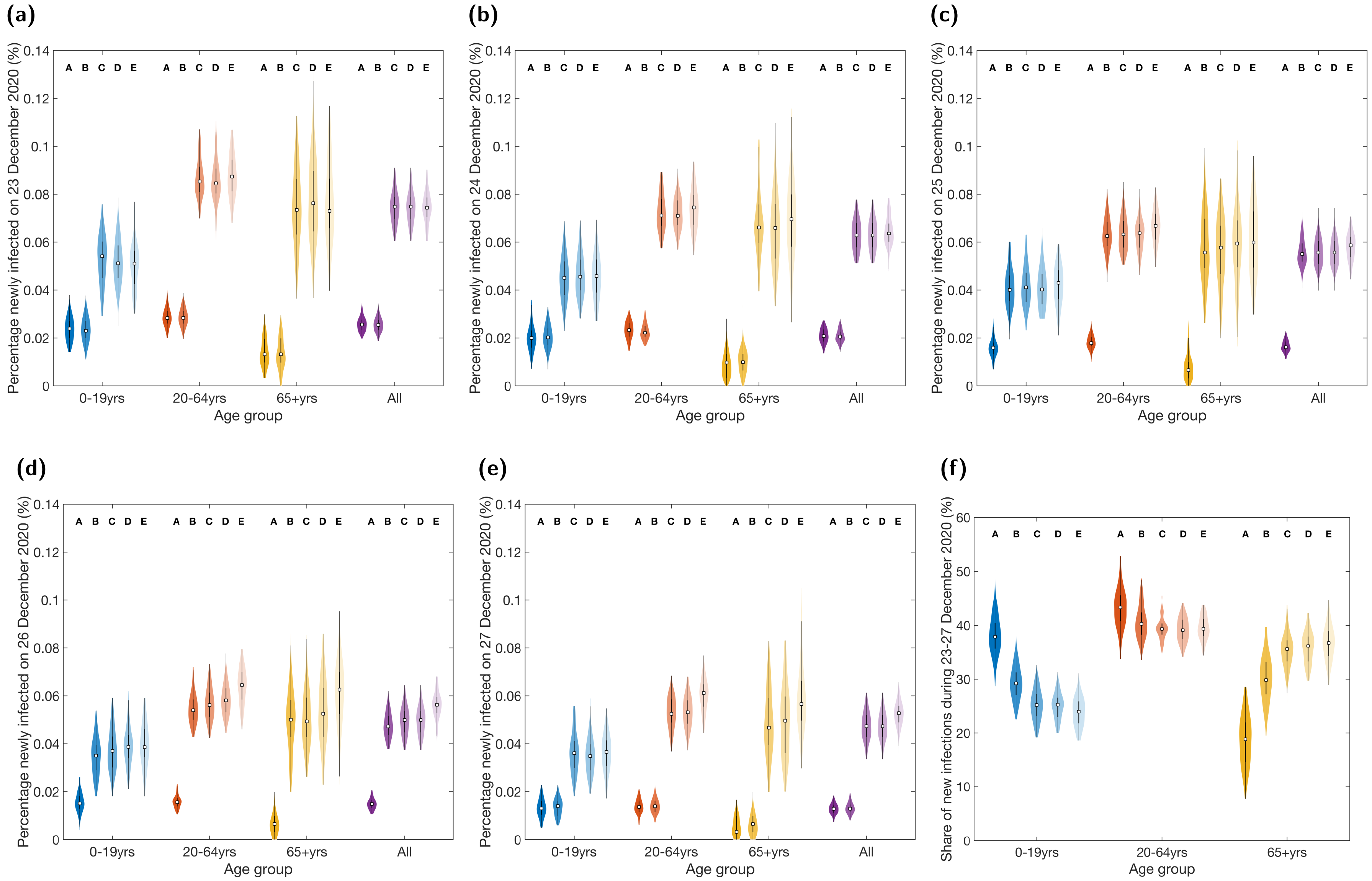


**Fig. S6: Distributions for the daily incidence and age-stratification of new infections between 23-27 December 2020 under each Christmas bubble scenario for the alternative analysis (with simulations beginning on 13 December 2022) combined with the lower adherence to testing, contact tracing and isolation measures assumption.** In these simulations, each household had a 30% probability of being adherent to the guidance on testing, contact tracing and isolation. Estimates produced from 100 realisations per scenario for daily incidence on: **(a)** 23 December 2020; **(b)** 24 December 2020; **(c)** 25 December 2020; **(d)** 26 December 2020; **(e)** 27 December 2020. Panel **(f)** displays the share of new infections between 23-27 December 2020 attributable to each of the three age groups. In each violin plot the white squares represent the medians and solid black lines correspond to the interquartile range. See Table S3 for median values and 95% prediction intervals. Labels A-E correspond to the distributions associated with Scenarios A-E. The intensity of shading of the violin plots also distinguishes between the scenarios; darkest for Scenario A to lightest for Scenario E. Scenario A: No change (support bubbles only); Scenario B: Short duration fixed exclusive bubbles, meet 25 and 26 December only; Scenario C: Fixed exclusive bubbles, meet every day 23-27 December; Scenario D: Fixed non-exclusive bubbles, meet every day 23-27 December; Scenario E: New household triplets meet each day between 23-27 December.

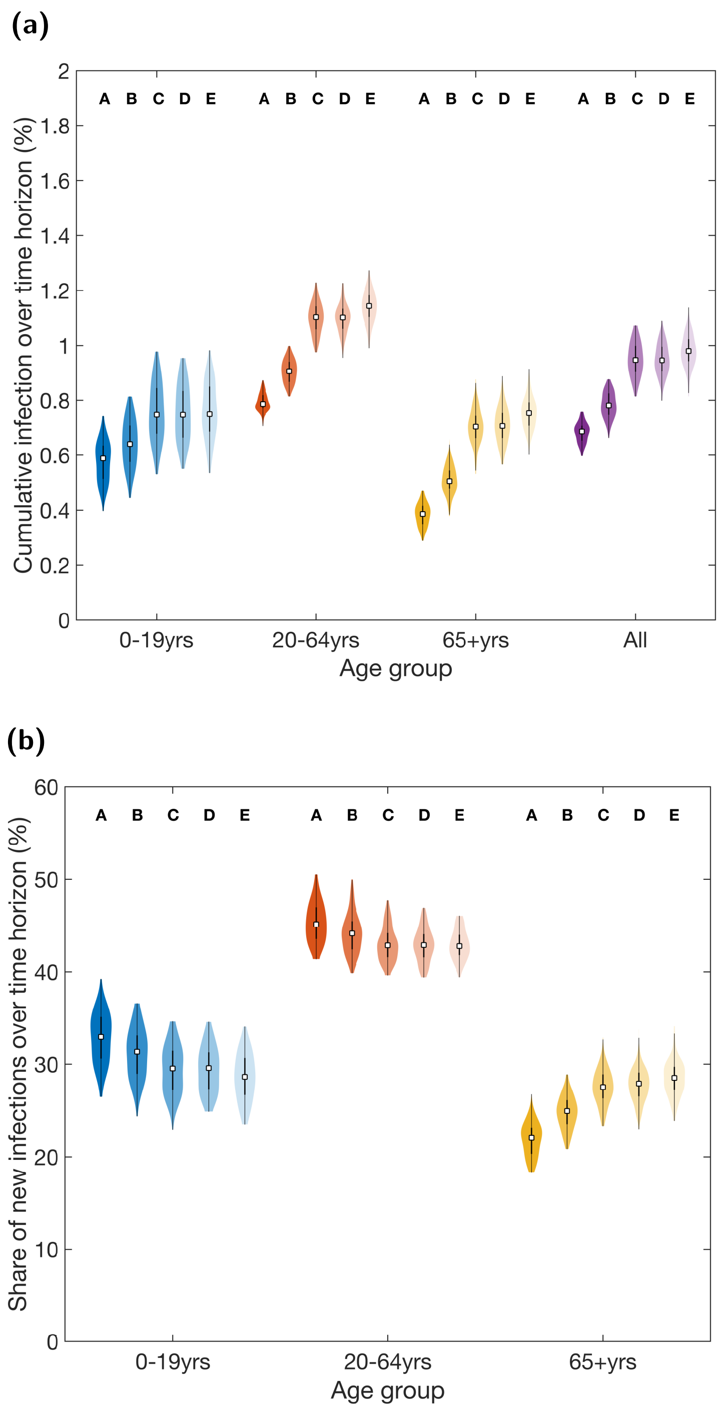


**Fig. S7: For the alternative analysis with simulations beginning on 13 December 2022 and lower adherence to testing, contact tracing and isolation measures, cumulative infection distributions for the entire 25 day time horizon, 13 December 2020 to 06 January 2021, under each Christmas bubble scenario.** In these simulations, each household had a 30% probability of being adherent to the guidance on testing, contact tracing and isolation. Estimates produced from 100 realisations per scenario. **(a)** The percentage of each age group newly infected during the time horizon. **(b)** The percentage of infections during the time horizon attributed to each age group. In each violin plot the white squares represent the medians and solid black lines correspond to the interquartile range. See Table S3 for median values and 95% prediction intervals. Labels A-E correspond to the distributions associated with Scenarios A-E. The intensity of shading of the violin plots also distinguishes between the scenarios; darkest for Scenario A to lightest for Scenario E. Scenario A: No change (support bubbles only); Scenario B: Short duration fixed exclusive bubbles, meet 25 and 26 December only; Scenario C: Fixed exclusive bubbles, meet every day 23-27 December; Scenario D: Fixed non-exclusive bubbles, meet every day 23-27 December; Scenario E: New household triplets meet each day between 23-27 December.

### Additional tables

**Table S1: Summary statistics for the epidemiological outcomes in our main analysis between 23-27 December 2020 under the five household bubbling scenarios.** For the 100 simulations performed per scenario, we report medians and in parentheses the 95% prediction intervals. We report estimates for each statistic to the following precision: percentage newly infected to 3 decimal places; percentage of those infected between 23-27 December 2020 attributable to each age group to the nearest integer.

| | | Scenario | | | | |
|---|---|---|---|---|---|---|
| **Statistic** | **Age group (years)** | **A** | **B** | **C** | **D** | **E** |
| **Percentage newly infected on 23 December 2020** | **0-19** | 0.028% (0.014%,0.039%) | 0.029% (0.019%,0.040%) | 0.061% (0.042%,0.085%) | 0.059% (0.041%,0.080%) | 0.059% (0.040%,0.083%) |
| | **20-64** | 0.047% (0.038%,0.058%) | 0.047% (0.038%,0.059%) | 0.106% (0.093%,0.125%) | 0.108% (0.092%,0.128%) | 0.108% (0.091%,0.129%) |
| | **65+** | 0.023% (0.007%,0.046%) | 0.023% (0.010%,0.046%) | 0.092% (0.063%,0.148%) | 0.096% (0.063%,0.130%) | 0.096% (0.060%,0.136%) |
| | **All** | 0.039% (0.030%,0.046%) | 0.039% (0.033%,0.048%) | 0.092% (0.077%,0.103%) | 0.092% (0.077%,0.103%) | 0.092% (0.077%,0.108%) |
| **Percentage newly infected on 24 December 2020** | **0-19** | 0.037% (0.022%,0.052%) | 0.037% (0.023%,0.049%) | 0.073% (0.049%,0.096%) | 0.074% (0.049%,0.099%) | 0.073% (0.051%,0.098%) |
| | **20-64** | 0.058% (0.049%,0.071%) | 0.059% (0.046%,0.072%) | 0.131% (0.115%,0.151%) | 0.129% (0.114%,0.148%) | 0.131% (0.116%,0.150%) |
| | **65+** | 0.030% (0.013%,0.047%) | 0.033% (0.013%,0.054%) | 0.118% (0.073%,0.157%) | 0.113% (0.076%,0.161%) | 0.116% (0.077%,0.152%) |
| | **All** | 0.048% (0.040%,0.056%) | 0.049% (0.041%,0.059%) | 0.111% (0.098%,0.124%) | 0.111% (0.098%,0.124%) | 0.112% (0.094%,0.126%) |
| **Percentage newly infected on 25 December 2020** | **0-19** | 0.044% (0.028%,0.058%) | 0.087% (0.066%,0.117%) | 0.090% (0.066%,0.113%) | 0.090% (0.066%,0.115%) | 0.089% (0.063%,0.119%) |
| | **20-64** | 0.072% (0.061%,0.083%) | 0.157% (0.138%,0.178%) | 0.157% (0.141%,0.176%) | 0.157% (0.139%,0.176%) | 0.160% (0.144%,0.175%) |
| | **65+** | 0.033% (0.017%,0.060%) | 0.132% (0.096%,0.173%) | 0.131% (0.093%,0.185%) | 0.136% (0.096%,0.171%) | 0.132% (0.099%,0.179%) |
| | **All** | 0.059% (0.051%,0.067%) | 0.132% (0.115%,0.149%) | 0.133% (0.118%,0.147%) | 0.133% (0.118%,0.147%) | 0.134% (0.120%,0.150%) |
| **Percentage newly infected on 26 December 2020** | **0-19** | 0.052% (0.032%,0.068%) | 0.103% (0.073%,0.132%) | 0.103% (0.075%,0.137%) | 0.102% (0.076%,0.134%) | 0.101% (0.072%,0.132%) |
| | **20-64** | 0.081% (0.068%,0.094%) | 0.176% (0.152%,0.197%) | 0.178% (0.155%,0.202%) | 0.178% (0.154%,0.201%) | 0.183% (0.159%,0.203%) |
| | **65+** | 0.040% (0.023%,0.057%) | 0.145% (0.110%,0.185%) | 0.150% (0.097%,0.192%) | 0.150% (0.106%,0.201%) | 0.151% (0.106%,0.195%) |
| | **All** | 0.067% (0.056%,0.077%) | 0.148% (0.133%,0.168%) | 0.151% (0.131%,0.171%) | 0.151% (0.131%,0.171%) | 0.154% (0.136%,0.171%) |
| **Percentage newly infected on 27 December 2020** | **0-19** | 0.052% (0.034%,0.073%) | 0.053% (0.037%,0.075%) | 0.095% (0.066%,0.130%) | 0.090% (0.065%,0.121%) | 0.094% (0.061%,0.125%) |
| | **20-64** | 0.081% (0.069%,0.092%) | 0.083% (0.070%,0.096%) | 0.157% (0.140%,0.177%) | 0.157% (0.136%,0.175%) | 0.164% (0.146%,0.179%) |
| | **65+** | 0.036% (0.013%,0.060%) | 0.036% (0.017%,0.060%) | 0.117% (0.090%,0.159%) | 0.123% (0.083%,0.182%) | 0.125% (0.089%,0.172%) |
| | **All** | 0.067% (0.058%,0.079%) | 0.069% (0.061%,0.080%) | 0.134% (0.118%,0.154%) | 0.134% (0.118%,0.154%) | 0.138% (0.121%,0.154%) |
| **Share of cumulative infections between 23-27 December 2020** | **0-19** | 30% (24%,34%) | 26% (21%,31%) | 24% (19%,28%) | 24% (20%,28%) | 23% (19%,27%) |
| | **20-64** | 47% (42%,53%) | 43% (40%,48%) | 41% (38%,45%) | 41% (39%,45%) | 42% (38%,46%) |
| | **65+** | 23% (19%,29%) | 31% (27%,35%) | 34% (31%,39%) | 35% (32%,39%) | 35% (30%,39%) |

**Table S2: Summary statistics for the epidemiological outcomes in our main analysis over the whole time horizon (23 December 2020 - 06 January 2021) under the five household bubbling scenarios.** For the 100 simulations performed per scenario, we report medians and in parentheses the 95% prediction intervals. We report estimates for each statistic to the following precision: cumulative infections to 2 decimal places; percentage of those infected stratified by age group to the nearest integer.

| | | Scenario | | | | |
|---|---|---|---|---|---|---|
| **Statistic** | **Age group (years)** | **A** | **B** | **C** | **D** | **E** |
| **Cumulative infection** | **0-19** | 0.49% (0.38%,0.58%) | 0.63% (0.50%,0.76%) | 0.78% (0.60%,0.95%) | 0.79% (0.61%,0.94%) | 0.80% (0.60%,0.98%) |
| | **20-64** | 0.71% (0.64%,0.76%) | 0.96% (0.89%,1.02%) | 1.23% (1.14%,1.32%) | 1.23% (1.12%,1.33%) | 1.26% (1.13%,1.34%) |
| | **65+** | 0.32% (0.26%,0.39%) | 0.58% (0.49%,0.67%) | 0.87% (0.73%,0.99%) | 0.89% (0.76%,1.01%) | 0.90% (0.75%,1.06%) |
| | **All** | 0.60% (0.53%,0.64%) | 0.82% (0.75%,0.89%) | 1.05% (0.95%,1.15%) | 1.05% (0.95%,1.15%) | 1.08% (0.94%,1.18%) |
| **Share of cumulative infections** | **0-19** | 32% (28%,37%) | 29% (25%,33%) | 27% (23%,32%) | 27% (23%,31%) | 27% (23%,31%) |
| | **20-64** | 47% (43%,51%) | 44% (41%,49%) | 43% (40%,46%) | 42% (39%,45%) | 43% (40%,46%) |
| | **65+** | 21% (17%,24%) | 27% (23%,30%) | 30% (27%,34%) | 31% (27%,34%) | 31% (27%,34%) |

**Table S3: Summary statistics for the epidemiological outcomes in our alternative analysis between 23-27 December 2020 under the five household bubbling scenarios**. For the 100 simulations performed per scenario, we report medians and in parentheses the 95% prediction intervals. We report estimates for each statistic to the following precision: percentage newly infected to 3 decimal places; percentage of those infected between 23-27 December 2020 attributable to each age group to the nearest integer.

| | | Scenario | | | | |
|---|---|---|---|---|---|---|
| **Statistic** | **Age group (years)** | **A** | **B** | **C** | **D** | **E** |
| **Percentage newly infected on 23 December 2020** | **0-19** | 0.022% (0.011%,0.033%) | 0.023% (0.013%,0.033%) | 0.037% (0.026%,0.049%) | 0.034% (0.022%,0.053%) | 0.036% (0.020%,0.053%) |
| | **20-64** | 0.026% (0.019%,0.035%) | 0.026% (0.018%,0.033%) | 0.054% (0.040%,0.068%) | 0.054% (0.040%,0.065%) | 0.053% (0.043%,0.065%) |
| | **65+** | 0.010% (0.000%,0.020%) | 0.010% (0.003%,0.020%) | 0.043% (0.017%,0.067%) | 0.043% (0.023%,0.066%) | 0.043% (0.017%,0.066%) |
| | **All** | 0.024% (0.017%,0.029%) | 0.023% (0.018%,0.029%) | 0.048% (0.037%,0.058%) | 0.048% (0.037%,0.058%) | 0.046% (0.038%,0.056%) |
| **Percentage newly infected on 24 December 2020** | **0-19** | 0.018% (0.010%,0.028%) | 0.018% (0.011%,0.030%) | 0.030% (0.017%,0.043%) | 0.028% (0.018%,0.044%) | 0.030% (0.018%,0.041%) |
| | **20-64** | 0.021% (0.015%,0.030%) | 0.021% (0.015%,0.029%) | 0.043% (0.036%,0.058%) | 0.043% (0.036%,0.055%) | 0.045% (0.033%,0.055%) |
| | **65+** | 0.007% (0.000%,0.020%) | 0.007% (0.000%,0.020%) | 0.033% (0.013%,0.060%) | 0.036% (0.020%,0.050%) | 0.033% (0.017%,0.054%) |
| | **All** | 0.019% (0.014%,0.024%) | 0.019% (0.015%,0.025%) | 0.038% (0.031%,0.049%) | 0.038% (0.031%,0.049%) | 0.039% (0.030%,0.046%) |
| **Percentage newly infected on 25 December 2020** | **0-19** | 0.015% (0.008%,0.026%) | 0.025% (0.013%,0.039%) | 0.027% (0.016%,0.038%) | 0.026% (0.016%,0.040%) | 0.025% (0.015%,0.038%) |
| | **20-64** | 0.016% (0.010%,0.025%) | 0.037% (0.026%,0.046%) | 0.036% (0.027%,0.045%) | 0.037% (0.028%,0.048%) | 0.038% (0.030%,0.047%) |
| | **65+** | 0.007% (0.000%,0.013%) | 0.027% (0.010%,0.053%) | 0.030% (0.010%,0.047%) | 0.030% (0.010%,0.053%) | 0.030% (0.010%,0.056%) |
| | **All** | 0.015% (0.010%,0.021%) | 0.032% (0.025%,0.041%) | 0.033% (0.025%,0.039%) | 0.033% (0.025%,0.039%) | 0.034% (0.027%,0.040%) |
| **Percentage newly infected on 26 December 2020** | **0-19** | 0.013% (0.006%,0.024%) | 0.022% (0.011%,0.032%) | 0.023% (0.013%,0.036%) | 0.023% (0.011%,0.036%) | 0.024% (0.013%,0.036%) |
| | **20-64** | 0.014% (0.009%,0.020%) | 0.031% (0.022%,0.040%) | 0.033% (0.024%,0.042%) | 0.033% (0.024%,0.044%) | 0.035% (0.026%,0.043%) |
| | **65+** | 0.003% (0.000%,0.017%) | 0.023% (0.007%,0.046%) | 0.026% (0.007%,0.047%) | 0.026% (0.007%,0.050%) | 0.027% (0.010%,0.047%) |
| | **All** | 0.013% (0.009%,0.017%) | 0.027% (0.021%,0.033%) | 0.029% (0.022%,0.037%) | 0.029% (0.022%,0.037%) | 0.031% (0.024%,0.038%) |
| **Percentage newly infected on 27 December 2020** | **0-19** | 0.011% (0.005%,0.018%) | 0.012% (0.006%,0.018%) | 0.021% (0.013%,0.034%) | 0.022% (0.012%,0.034%) | 0.022% (0.013%,0.036%) |
| | **20-64** | 0.012% (0.007%,0.016%) | 0.012% (0.007%,0.016%) | 0.029% (0.021%,0.037%) | 0.030% (0.020%,0.038%) | 0.033% (0.021%,0.044%) |
| | **65+** | 0.003% (0.000%,0.013%) | 0.003% (0.000%,0.013%) | 0.022% (0.007%,0.037%) | 0.023% (0.007%,0.040%) | 0.027% (0.003%,0.047%) |
| | **All** | 0.011% (0.007%,0.015%) | 0.011% (0.007%,0.015%) | 0.026% (0.020%,0.033%) | 0.026% (0.020%,0.033%) | 0.028% (0.020%,0.037%) |
| **Share of cumulative infections between 23-27 December 2020** | **0-19** | 40% (30%,48%) | 33% (25%,40%) | 28% (23%,33%) | 27% (21%,33%) | 27% (20%,33%) |
| | **20-64** | 45% (37%,53%) | 42% (37%,51%) | 40% (36%,46%) | 41% (35%,47%) | 40% (36%,46%) |
| | **65+** | 16% (8%,24%)' } | 24% (16%,36%) | 31% (23%,37%) | 32% (24%,40%) | 32% (24%,38%) |

**Table S4: Summary statistics for the epidemiological outcomes in our alternative analysis over the whole time horizon (13 December 2020 - 06 January 2021) under the five household bubbling scenarios.** For the 100 simulations performed per scenario, we report medians and in parentheses the 95% prediction intervals. We report estimates for each statistic to the following precision: cumulative infections to 2 decimal places; percentage of those infected stratified by age group to the nearest integer.

| | | Scenario | | | | |
|---|---|---|---|---|---|---|
| **Statistic** | **Age group (years)** | **A** | **B** | **C** | **D** | **E** |
| **Cumulative infection** | **0-19** | 0.54% (0.42%,0.64%) | 0.57% (0.43%,0.70%) | 0.62% (0.48%,0.75%) | 0.62% (0.46%,0.76%) | 0.63% (0.48%,0.77%) |
| | **20-64** | 0.75% (0.69%,0.81%) | 0.81% (0.74%,0.86%) | 0.90% (0.82%,0.96%) | 0.90% (0.82%,0.97%) | 0.91% (0.82%,0.97%) |
| | **65+** | 0.33% (0.27%,0.42%) | 0.38% (0.31%,0.46%) | 0.48% (0.39%,0.56%) | 0.49% (0.39%,0.59%) | 0.50% (0.41%,0.59%) |
| | **All** | 0.65% (0.57%,0.69%) | 0.69% (0.63%,0.75%) | 0.77% (0.68%,0.85%) | 0.77% (0.68%,0.85%) | 0.78% (0.68%,0.85%) |
| **Share of cumulative infections** | **0-19** | 33% (29%,38%) | 33% (27%,36%) | 31% (27%,35%) | 31% (26%,35%) | 31% (27%,35%) |
| | **20-64** | 46% (43%,50%) | 46% (41%,50%) | 45% (41%,48%) | 45% (41%,49%) | 44% (41%,49%) |
| | **65+** | 20% (17%,25%) | 22% (18%,26%) | 24% (20%,27%) | 24% (21%,28%) | 24% (21%,28%) |

**Table S5: Summary statistics for the epidemiological outcomes in our main analysis between 23-27 December 2020 under the five household bubbling scenarios and against a backdrop of a lower adherence to testing, contact tracing and isolation measures (each household having a 30% probability of being adherent).** For the 100 simulations performed per scenario, we report medians and in parentheses the 95% prediction intervals. We report estimates for each statistic to the following precision: percentage newly infected to 3 decimal places; percentage of those infected between 23-27 December 2020 attributable to each age group to the nearest integer.

| | | Scenario | | | | |
|---|---|---|---|---|---|---|
| **Statistic** | **Age group (years)** | **A** | **B** | **C** | **D** | **E** |
| **Percentage newly infected on 23 December 2020** | **0-19** | 0.029% (0.017%,0.044%) | 0.030% (0.020%,0.042%) | 0.062% (0.041%,0.089%) | 0.063% (0.045%,0.084%) | 0.064% (0.045%,0.088%) |
| | **20-64** | 0.047% (0.037%,0.058%) | 0.049% (0.040%,0.060%) | 0.116% (0.101%,0.132%) | 0.118% (0.100%,0.135%) | 0.117% (0.100%,0.135%) |
| | **65+** | 0.026% (0.007%,0.046%) | 0.026% (0.010%,0.046%) | 0.103% (0.070%,0.149%) | 0.106% (0.066%,0.141%) | 0.102% (0.060%,0.142%) |
| | **All** | 0.040% (0.031%,0.048%) | 0.040% (0.033%,0.047%) | 0.099% (0.084%,0.112%) | 0.099% (0.084%,0.112%) | 0.098% (0.087%,0.114%) |
| **Percentage newly infected on 24 December 2020** | **0-19** | 0.036% (0.020%,0.050%) | 0.036% (0.022%,0.056%) | 0.080% (0.059%,0.105%) | 0.083% (0.058%,0.104%) | 0.079% (0.056%,0.111%) |
| | **20-64** | 0.060% (0.050%,0.071%) | 0.060% (0.048%,0.071%) | 0.145% (0.127%,0.163%) | 0.145% (0.129%,0.165%) | 0.149% (0.130%,0.167%) |
| | **65+** | 0.033% (0.013%,0.053%) | 0.033% (0.013%,0.064%) | 0.128% (0.082%,0.162%) | 0.128% (0.089%,0.183%) | 0.133% (0.093%,0.175%) |
| | **All** | 0.050% (0.041%,0.058%) | 0.050% (0.040%,0.060%) | 0.124% (0.111%,0.140%) | 0.124% (0.111%,0.140%) | 0.125% (0.109%,0.146%) |
| **Percentage newly infected on 25 December 2020** | **0-19** | 0.044% (0.032%,0.063%) | 0.100% (0.070%,0.126%) | 0.100% (0.070%,0.134%) | 0.100% (0.070%,0.128%) | 0.100% (0.071%,0.131%) |
| | **20-64** | 0.073% (0.063%,0.086%) | 0.180% (0.162%,0.205%) | 0.181% (0.160%,0.202%) | 0.181% (0.161%,0.207%) | 0.185% (0.164%,0.208%) |
| | **65+** | 0.040% (0.020%,0.063%) | 0.159% (0.122%,0.204%) | 0.163% (0.109%,0.205%) | 0.166% (0.122%,0.212%) | 0.166% (0.116%,0.215%) |
| | **All** | 0.061% (0.051%,0.072%) | 0.154% (0.138%,0.169%) | 0.154% (0.136%,0.169%) | 0.154% (0.136%,0.169%) | 0.156% (0.139%,0.176%) |
| **Percentage newly infected on 26 December 2020** | **0-19** | 0.054% (0.035%,0.075%) | 0.119% (0.082%,0.147%) | 0.118% (0.083%,0.148%) | 0.120% (0.087%,0.152%) | 0.120% (0.088%,0.149%) |
| | **20-64** | 0.083% (0.070%,0.101%) | 0.205% (0.186%,0.231%) | 0.207% (0.180%,0.234%) | 0.210% (0.180%,0.230%) | 0.217% (0.193%,0.243%) |
| | **65+** | 0.043% (0.016%,0.072%) | 0.182% (0.132%,0.238%) | 0.183% (0.136%,0.231%) | 0.188% (0.139%,0.233%) | 0.192% (0.139%,0.238%) |
| | **All** | 0.069% (0.060%,0.081%) | 0.176% (0.158%,0.193%) | 0.177% (0.155%,0.199%) | 0.177% (0.155%,0.199%) | 0.183% (0.165%,0.206%) |
| **Percentage newly infected on 27 December 2020** | **0-19** | 0.055% (0.032%,0.075%) | 0.056% (0.040%,0.078%) | 0.121% (0.083%,0.148%) | 0.124% (0.089%,0.159%) | 0.123% (0.090%,0.159%) |
| | **20-64** | 0.084% (0.070%,0.098%) | 0.087% (0.071%,0.104%) | 0.207% (0.190%,0.237%) | 0.209% (0.184%,0.234%) | 0.223% (0.192%,0.248%) |
| | **65+** | 0.040% (0.020%,0.067%) | 0.043% (0.026%,0.073%) | 0.179% (0.142%,0.234%) | 0.178% (0.127%,0.232%) | 0.202% (0.154%,0.253%) |
| | **All** | 0.071% (0.059%,0.083%) | 0.073% (0.062%,0.087%) | 0.178% (0.157%,0.200%) | 0.178% (0.157%,0.200%) | 0.189% (0.164%,0.209%) |
| **Share of cumulative infections between 23-27 December 2020** | **0-19** | 29% (23%,34%) | 25% (20%,29%) | 23% (19%,27%) | 23% (19%,27%) | 22% (18%,27%) |
| | **20-64** | 47% (43%,52%) | 43% (38%,47%) | 41% (38%,44%) | 41% (37%,44%) | 41% (38%,44%) |
| | **65+** | 24% (18%,30%) | 32% (28%,36%) | 36% (32%,39%) | 36% (33%,41%) | 37% (33%,40%) |

**Table S6: Summary statistics for the epidemiological outcomes from our adherence sensitivity analysis, with a lower adherence to testing, contact tracing and isolation measures (each household having a 30% probability of being adherent).** Outcomes were calculated over the whole time horizon (23 December 2020 - 06 January 2021) for each of the five household bubbling scenarios. For the 100 simulations performed per scenario, we report medians and in parentheses the 95% prediction intervals. We report estimates for each statistic to the following precision: cumulative infections to 2 decimal places; percentage of those infected stratified by age group to the nearest integer.

| | | Scenario | | | | |
|---|---|---|---|---|---|---|
| **Statistic** | **Age group (years)** | **A** | **B** | **C** | **D** | **E** |
| **Cumulative infection** | **0-19** | 0.53% (0.41%,0.63%) | 0.70% (0.52%,0.83%) | 0.90% (0.70%,1.07%) | 0.91% (0.70%,1.11%) | 0.92% (0.72%,1.16%) |
| | **20-64** | 0.74% (0.68%,0.80%) | 1.06% (0.98%,1.14%) | 1.42% (1.31%,1.55%) | 1.42% (1.31%,1.54%) | 1.49% (1.36%,1.58%) |
| | **65+** | 0.36% (0.29%,0.43%) | 0.70% (0.60%,0.85%) | 1.11% (0.92%,1.26%) | 1.11% (0.97%,1.26%) | 1.17% (0.98%,1.32%) |
| | **All** | 0.64% (0.56%,0.70%) | 0.91% (0.82%,0.99%) | 1.22% (1.10%,1.32%) | 1.23% (1.09%,1.33%) | 1.27% (1.13%,1.37%) |
| **Share of cumulative infections** | **0-19** | 32% (27%,36%) | 28% (24%,32%) | 26% (23%,30%) | 26% (23%,30%) | 26% (22%,31%) |
| | **20-64** | 46% (42%,50%) | 43% (40%,47%) | 42% (39%,45%) | 42% (38%,44%) | 42% (38%,45%) |
| | **65+** | 22% (19%,26%) | 29% (25%,32%) | 32% (29%,35%) | 32% (29%,36%) | 33% (29%,36%) |

**Table S7: Summary statistics for the epidemiological outcomes in our alternative analysis between 23-27 December 2020 under the five household bubbling scenarios and against a backdrop of a lower adherence to testing, contact tracing and isolation measures (each household having a 30% probability of being adherent).** For the 100 simulations performed per scenario, we report medians and in parentheses the 95% prediction intervals. We report estimates for each statistic to the following precision: percentage newly infected to 3 decimal places; percentage of those infected between 23-27 December 2020 attributable to each age group to the nearest integer.

| | | Scenario | | | | |
|---|---|---|---|---|---|---|
| **Statistic** | **Age group (years)** | **A** | **B** | **C** | **D** | **E** |
| **Percentage newly infected on 23 December 2020** | **0-19** | 0.024% (0.016%,0.034%) | 0.023% (0.014%,0.034%) | 0.054% (0.033%,0.071%) | 0.051% (0.035%,0.072%) | 0.051% (0.033%,0.073%) |
| | **20-64** | 0.028% (0.021%,0.037%) | 0.028% (0.021%,0.036%) | 0.085% (0.071%,0.105%) | 0.085% (0.067%,0.105%) | 0.087% (0.072%,0.106%) |
| | **65+** | 0.013% (0.003%,0.027%) | 0.013% (0.000%,0.027%) | 0.074% (0.047%,0.106%) | 0.076% (0.047%,0.113%) | 0.073% (0.046%,0.109%) |
| | **All** | 0.026% (0.021%,0.032%) | 0.026% (0.019%,0.032%) | 0.075% (0.061%,0.089%) | 0.075% (0.061%,0.089%) | 0.074% (0.061%,0.089%) |
| **Percentage newly infected on 24 December 2020** | **0-19** | 0.020% (0.010%,0.033%) | 0.020% (0.011%,0.030%) | 0.045% (0.030%,0.064%) | 0.046% (0.032%,0.066%) | 0.046% (0.029%,0.065%) |
| | **20-64** | 0.023% (0.016%,0.029%) | 0.022% (0.017%,0.031%) | 0.071% (0.058%,0.089%) | 0.071% (0.059%,0.085%) | 0.075% (0.060%,0.090%) |
| | **65+** | 0.010% (0.000%,0.023%) | 0.010% (0.000%,0.023%) | 0.066% (0.040%,0.100%) | 0.066% (0.036%,0.097%) | 0.070% (0.033%,0.110%) |
| | **All** | 0.021% (0.014%,0.027%) | 0.021% (0.016%,0.026%) | 0.063% (0.052%,0.077%) | 0.063% (0.052%,0.077%) | 0.064% (0.051%,0.078%) |
| **Percentage newly infected on 25 December 2020** | **0-19** | 0.016% (0.008%,0.024%) | 0.040% (0.027%,0.058%) | 0.041% (0.027%,0.061%) | 0.040% (0.028%,0.058%) | 0.043% (0.027%,0.057%) |
| | **20-64** | 0.018% (0.012%,0.025%) | 0.063% (0.051%,0.077%) | 0.063% (0.052%,0.078%) | 0.064% (0.050%,0.076%) | 0.067% (0.052%,0.079%) |
| | **65+** | 0.007% (0.000%,0.024%) | 0.056% (0.030%,0.089%) | 0.058% (0.030%,0.082%) | 0.059% (0.020%,0.097%) | 0.060% (0.033%,0.089%) |
| | **All** | 0.016% (0.012%,0.022%) | 0.055% (0.045%,0.068%) | 0.056% (0.047%,0.066%) | 0.056% (0.047%,0.066%) | 0.059% (0.046%,0.069%) |
| **Percentage newly infected on 26 December 2020** | **0-19** | 0.015% (0.008%,0.026%) | 0.035% (0.021%,0.052%) | 0.037% (0.021%,0.056%) | 0.039% (0.024%,0.056%) | 0.039% (0.022%,0.058%) |
| | **20-64** | 0.016% (0.012%,0.022%) | 0.054% (0.044%,0.070%) | 0.056% (0.045%,0.071%) | 0.058% (0.048%,0.071%) | 0.065% (0.051%,0.078%) |
| | **65+** | 0.007% (0.000%,0.017%) | 0.050% (0.023%,0.077%) | 0.049% (0.030%,0.076%) | 0.053% (0.027%,0.083%) | 0.063% (0.036%,0.086%) |
| | **All** | 0.015% (0.011%,0.020%) | 0.047% (0.039%,0.059%) | 0.050% (0.040%,0.063%) | 0.050% (0.040%,0.063%) | 0.056% (0.045%,0.067%) |
| **Percentage newly infected on 27 December 2020** | **0-19** | 0.013% (0.006%,0.022%) | 0.014% (0.006%,0.022%) | 0.036% (0.023%,0.055%) | 0.035% (0.020%,0.053%) | 0.037% (0.018%,0.051%) |
| | **20-64** | 0.014% (0.007%,0.020%) | 0.014% (0.008%,0.021%) | 0.053% (0.041%,0.064%) | 0.053% (0.042%,0.067%) | 0.061% (0.048%,0.073%) |
| | **65+** | 0.003% (0.000%,0.013%) | 0.007% (0.000%,0.017%) | 0.047% (0.026%,0.077%) | 0.050% (0.023%,0.080%) | 0.057% (0.033%,0.096%) |
| | **All** | 0.013% (0.008%,0.017%) | 0.013% (0.009%,0.019%) | 0.047% (0.034%,0.059%) | 0.047% (0.034%,0.059%) | 0.053% (0.042%,0.063%) |
| **Share of cumulative infections between 23-27 December 2020** | **0-19** | 38% (31%,46%) | 29% (24%,35%) | 25% (20%,30%) | 25% (20%,29%) | 24% (19%,29%) |
| | **20-64** | 43% (35%,52%) | 40% (35%,47%) | 39% (36%,45%) | 39% (35%,43%) | 39% (36%,43%) |
| | **65+** | 19% (10%,28%) | 30% (22%,39%) | 36% (30%,42%) | 36% (30%,41%) | 37% (29%,43%) |

**Table S8: Summary statistics for the epidemiological outcomes from our adherence sensitivity analysis, with a lower adherence to testing, contact tracing and isolation measures (each household having a 30% probability of being adherent).** Outcomes were calculated over the extended time horizon (13 December 2020 - 06 January 2021) for each of the five household bubbling scenarios. For the 100 simulations performed per scenario, we report medians and in parentheses the 95% prediction intervals. We report estimates for each statistic to the following precision: cumulative infections to 2 decimal places; percentage of those infected stratified by age group to the nearest integer.

| | | Scenario | | | | |
|---|---|---|---|---|---|---|
| **Statistic** | **Age group (years)** | **A** | **B** | **C** | **D** | **E** |
| **Cumulative infection** | **0-19** | 0.59% (0.45%,0.71%) | 0.64% (0.48%,0.80%) | 0.75% (0.57%,0.95%) | 0.75% (0.58%,0.93%) | 0.75% (0.56%,0.93%) |
| | **20-64** | 0.79% (0.73%,0.86%) | 0.91% (0.83%,0.98%) | 1.10% (0.99%,1.21%) | 1.10% (0.99%,1.21%) | 1.14% (1.02%,1.24%) |
| | **65+** | 0.39% (0.31%,0.46%) | 0.50% (0.41%,0.60%) | 0.70% (0.57%,0.82%) | 0.71% (0.58%,0.84%) | 0.75% (0.65%,0.88%) |
| | **All** | 0.69% (0.61%,0.75%) | 0.78% (0.70%,0.86%) | 0.95% (0.83%,1.06%) | 0.95% (0.85%,1.06%) | 0.98% (0.85%,1.09%) |
| **Share of cumulative infections** | **0-19** | 33% (28%,37%) | 31% (26%,36%) | 30% (25%,34%) | 30% (25%,34%) | 29% (24%,33%) |
| | **20-64** | 45% (42%,50%) | 44% (41%,49%) | 43% (40%,47%) | 43% (40%,46%) | 43% (40%,46%) |
| | **65+** | 22% (19%,25%) | 25% (21%,29%) | 28% (24%,31%) | 28% (24%,31%) | 29% (25%,33%) |
